# Effectiveness of a telerehabilitation program for COVID-19 survivors (TERECO) on exercise capacity, pulmonary function, lower limb muscle strength, and quality of life: a randomized controlled trial

**DOI:** 10.1101/2021.03.08.21253007

**Authors:** Jian’an Li, Wenguang Xia, Chao Zhan, Shouguo Liu, Zhifei Yin, Jiayue Wang, Yufei Chong, Chanjuan Zheng, Xiaoming Fang, Wei Cheng, Jan D. Reinhardt

## Abstract

**Objectives:** To investigate superiority of a telerehabilitation program for Covid-19 (TERECO) over no rehabilitation with regard to functional exercise capacity, lower-limb muscle strength (LMS), pulmonary function, health-related quality of life (HRQOL), and perceived dyspnoea.

**Design:** Parallel-group randomised controlled-trial with 1:1 block-randomisation.

**Setting:** Three major hospitals from Jiangsu and Hubei provinces, China.

**Participants:** 120 Covid-19 survivors with modified Medical Research Council (mMRC) dyspnoea score of 2-3 who had been discharged from hospital were randomised. 61 were allocated to the control group and 59 to the TERECO group.

**Intervention:** The control group received educational instructions. The TERECO group participated in a 6-week home-based, pulmonary rehabilitation program delivered via smartphone and monitored with chest-worn heart rate telemetry. Exercise types comprised breathing control and thoracic expansion, aerobic exercise, and LMS exercise.

**Outcomes:** Primary outcome was 6-minute walking distance (6MWD) in metres. Secondary outcomes were LMS measured as squat time in seconds; pulmonary function assessed by spirometry with parameters being forced expiratory volume in 1 second (FEV1), forced vital capacity (FVC), FEV1/FVC, maximum voluntary ventilation (MVV), and peak expiratory flow; HRQOL measured with SF-12 physical component score (PCS) and mental component score (MCS); and mMRC dyspnoea, favourable outcome (no dyspnoea). Outcomes were assessed at 6 weeks (post-treatment) and 28 weeks (follow-up).

**Results:** 120 patients were randomised, 15 (12.5%) were lost to follow-up at study endpoint. No serious adverse events occurred. 38 participants in the TERECO group complied with the exercise protocol (64.41% of randomized). The adjusted between-group difference in change in 6MWD from baseline was 65.45 metres (95% CI 43.8-87.1, p<0.001) at post-treatment and 68.62 metres (95% CI 46.39-90.85, p<0.001) at follow-up. Treatment effects for LMS were 20.12 seconds (95% CI 12.34-27.9, p<0.001) post-treatment and 22.23 seconds (95% CI 14.24-30.21, p<0.001) at follow-up. No group differences were found for lung function apart from post-treatment MVV (10.57 litres/minute, 95% CI 0.26-17.88, p=0.005). Increase in SF-12 PCS was greater in the TERECO group with treatment effects estimated as 3.79 (95% CI 1.24-6.35, p=0.004) at post-treatment and 2.69 (95% CI 0.06-5.32, p=0.045) at follow-up. No significant between-group differences were found for improvements in SF-12 MCS. At post-treatment 90.4% endorsed a favourable outcome for mMRC dyspnoea in the TERECO group vs. 61.7% in control (adjusted RR 1.46, 1.17-1.82, p=0.001).

**Conclusions:** This trial demonstrated superiority of TERECO over no rehabilitation for 6MWD, LMS, and SF-12 PCS. We found no persistent effects on pulmonary function, SF-12 MCS, and perceived dyspnoea.

**Trial registration:** Chinese Clinical Trial Registry: ChiCTR2000031834, 11 Apr 2020, URL: http://www.chictr.org.cn/showproj.aspx?proj=52216

**KEY POINTS:** *What is already known:* Many Covid-19 survivors discharged from hospital have reduced exercise capacity, impaired pulmonary function, muscle weakness, and reduced quality of life, all of which might be addressed with pulmonary rehabilitation. However, evidence on effective pulmonary rehabilitation measures for this population is currently lacking. As delivery of conventional rehabilitation services is furthermore limited due to pandemic control measures, telerehabilitation programs represent a possible alternative.

*What the study adds:* We developed a telerehabilitation program for Covid-19 survivors (TERECO program) that is delivered via smartphone and can be carried out at home. Our study suggests that TERECO was safe and participants of the TERECO program had improved exercise capacity, lower-limb muscle strength, and physical quality of life. No relevant group differences were found for lung function, self-reported breathlessness, and mental quality of life. The TERECO program is inexpensive and could be implemented on a large scale to improve physical health of Covid-19 survivors after discharge from hospital.

## INTRODUCTION

After discharge from acute care, many survivors of coronavirus disease-2019 (Covid-19) experience ongoing symptoms^1 2^, impairment of pulmonary function^2–4^, decreased exercise capacity^2 4^, reduced muscle strength^5^, activity limitations^6 7^, anxiety^7^, depression and neurocognitive dysfunction^8^, and reduced quality of life^2^. Problems may persist for at least six months^2^. This indicates the need for the provision of rehabilitation services that can decrease the burden on patients and the health system^9^. Pulmonary rehabilitation measures with demonstrated effectiveness in chronic obstructive lung disease (COPD)^10^ and, with low-certainty evidence (one trial), severe acute respiratory syndrome (SARS)^11^ are obvious candidates.

Evidence on the effectiveness of such programs in Covid-19 survivors is, however, lacking to date^12^. Moreover, delivery of conventional in- or outpatient rehabilitation is complicated through diminished capacity in post-acute care as well as clinical and public health measures imposed to reduce the risk of viral transmission^13^. Telerehabilitation programs provide a viable alternative that could be superior to no rehabilitation (low-certainty evidence) and as effective as conventional rehabilitation (moderate-certainty evidence)^14 15^.

We investigated the possible superiority of a telerehabilitation program for Covid-19 (TERECO) over a control group that did not receive rehabilitation with regard to functional exercise capacity, lower-limb muscle strength, pulmonary function, perceived dyspnoea, and health-related quality of life in COVID-19 survivors who had been discharged to their homes following acute inpatient treatment. We further report on the occurrence of adverse events.

## METHODS

### Study design

This study is a multi-centre, parallel-group randomised controlled trial. It was registered at the Chinese Clinical Trial Registry on 11 Apr 2020 (ChiCTR2000031834). Ethical approval was first received from the Institutional Review Board (IRB) of the First Affiliated Hospital of Nanjing Medical University/Jiangsu Province Hospital (2020-SR-171, 9 April 2020) and then subsequently from the IRBs of Hubei Province Hospital of Integrated Chinese and Western Medicine (2020016, 14 April 2020), and Huangshi Hospital of Chinese Medicine (HSZYPJ-2020-026-01, 20 April 2020). The original protocol for this study is available from (URL: http://idmr.scu.edu.cn/info.htm?id=1841614474692833).

### Recruitment

Three centres from Jiangsu (The First Affiliated Hospital of Nanjing Medical University), Hubei Wuhan (Hubei Province Hospital of Integrated Chinese and Western Medicine), and Hubei Huangshi (Hubei Huangshi Hospital of Chinese Medicine) recruited patients recovering from COVID-19. Contact details of possible candidates were extracted from hospital records. The first contact was made by telephone. If the candidates expressed interest, an appointment for a baseline visit was made and informed written consent was obtained from all study participants at this baseline visit.

### Eligibility criteria

Hospital records were pre-screened for eligibility and further assessment of eligibility of possible candidates was performed at the baseline visit. Inclusion criteria were as follows: (i) laboratory confirmed SARS-2 infection, clinically diagnosed with COVID-19, having received inpatient treatment for it and having been discharged from one of the participating hospitals; (ii) aged 18-75 years; (iii) ownership of and ability to use a smartphone either independently or with support from family members; (iv) modified British Medical Research Council (mMRC) dyspnoea^16^ score of 2-3, indicating moderate dyspnoea symptoms. Participants were excluded under the following conditions: (i) resting heart rate of over 100 bpm as measured at the baseline visit; (ii) taking medication that can affect cardiopulmonary function and heart rate such as trimetazidine, bronchodilators, or β-blockers; (iii) medical history of cerebrovascular or cardiological condition within the past 12 months, severe kidney disease ( ≥ stage 3, eGFR<60ml/min), other severe organic disease, uncontrolled hypertension (resting BP≥160/100mmHg), uncontrolled diabetes (random blood glucose>16.7 mmol/l, HbA1C>7.0%); (iv) received intra-articular drug injection or surgical treatment of lower extremities within the past 6 months; (v) unable to walk independently with assistive device; (vi) unable or unwilling to collaborate with assessments; (vii) enrolled in or participated in other trials within the past 3 months; (viii) having severe cognitive or mental disorder or history of substance abuse; (ix) enrolment in any other rehabilitation program.

### Random sequence generation and allocation concealment

Permutated allocation sequences for 1:1 block randomisation (block size 10-14) stratified by hospital were computer-generated by an independent statistician. Allocation was concealed by central randomisation and only revealed after baseline assessment through a call to the study centre.

### Blinding

Participants and therapists could not be blinded. Assessors were blinded to group allocation by the following procedure: baseline visits of each potential study participant involved one assessor (rehabilitation doctor) and one independent allocator (therapist). Once a patient was deemed eligible and had provided informed consent, assessors conducted the baseline measurements. Assessors then left the study site and allocators contacted the study centre in the presence of the patient to reveal allocation. Patients and therapists were requested to not disclose allocation to the assessors at any time during the study.

### Interventions

#### Control group

Participants in the control group received 10-minute standardized educational instructions from physiotherapists and an information sheet containing these instructions in written form. They were advised to maintain normal daily activities, avoid excessive bed rest and immobilisation, take part in moderate physical activities such as housework, adhere to a healthy diet and get 6-8 hours of sleep per day. Patients also received advice on basic hand hygiene, physical distancing, and the appropriate use of personal protective equipment such as facemasks.

#### TERECO group

In addition to the instructions and information sheets received by the control group, the TERECO group took part in a 6-week remotely-monitored home exercise program delivered through a telerehabilitation smartphone application called RehabApp (R PLUS HEALTH, Version 1.1.0, Recovery Plus Inc., Chengdu, China). Participants received a heart rate (HR) telemetry device (Recovery Plus H1, Empsun Medical Co., Chengdu, China) to be worn on the chest during exercise. The HR telemetry device was connected to RehabApp through the internet. Physiotherapists with at least 5 years experience in rehabilitation regularly reviewed data on exercise duration and HR data. Patient feedback on the exercise program was also collected through RehabApp. In addition, consultations via mobile phone or WeChat voice calls were carried out every week. Issues raised by participants related to the exercise program were discussed and the exercise program was adjusted if necessary.

The duration of the exercise program was 40-60 minutes per session, with 3-4 sessions per week, for a total of 6 weeks. Initial exercise types and intensity were determined by the physical therapists based on baseline assessment and in accordance with the American College of Sports Medicine’s guidelines for exercise preparticipation^17^.

The telerehabilitation exercise program included (i) breathing control and thoracic expansion (BCTE), (ii) aerobic exercise, and (iii) lower-limb muscle strength (LMS) exercises specified in a three-tiered exercise plan as shown in Table 1. If patients met the target criteria for a tier 1 exercise during a particular week and did not report any adverse events, they could advance to tier 2 for this exercise type and so on. (i) BCTE included one diaphragmatic breathing exercise and several exercises aimed at improving thoracic muscle strength and chest mobility. (ii) Aerobic exercise was performed by brisk walking or running outside near the home or on a treadmill at home (if available to the participant) with a 3-minute warm-up and 3-minute stretching cool-down. The target heart rate (THR) for aerobic exercise was calculated with the Karvonen formula^18^, with exercise intensity scheduled to increase from 30-40% of heart rate reserve (HRR) at the beginning of the exercise program to 40-60% of HRR in the final two weeks of the program. Apart from monitoring the actual heart rate, patients were asked to report their rate of perceived exertion (RPE) after the aerobic exercises according to Borg’s scale^19 20^. When this RPE was greater than 14, the current intensity was considered unsuitable and the THR for a participant was lowered. When the reported RPE was below 11, participants were requested to increase their aerobic exercise intensity. When the reported RPE ranged between 11 and 14, participants repeated the exercise at the same intensity and moved to the next tier after 2 weeks. (iii) LMS exercise comprised weight-bearing and resistance exercises^21^ and gradually moved from exercises performed in a sitting position such as seated leg lifting to exercises executed in standing position such as squats and side lunges.

**Table 1:**
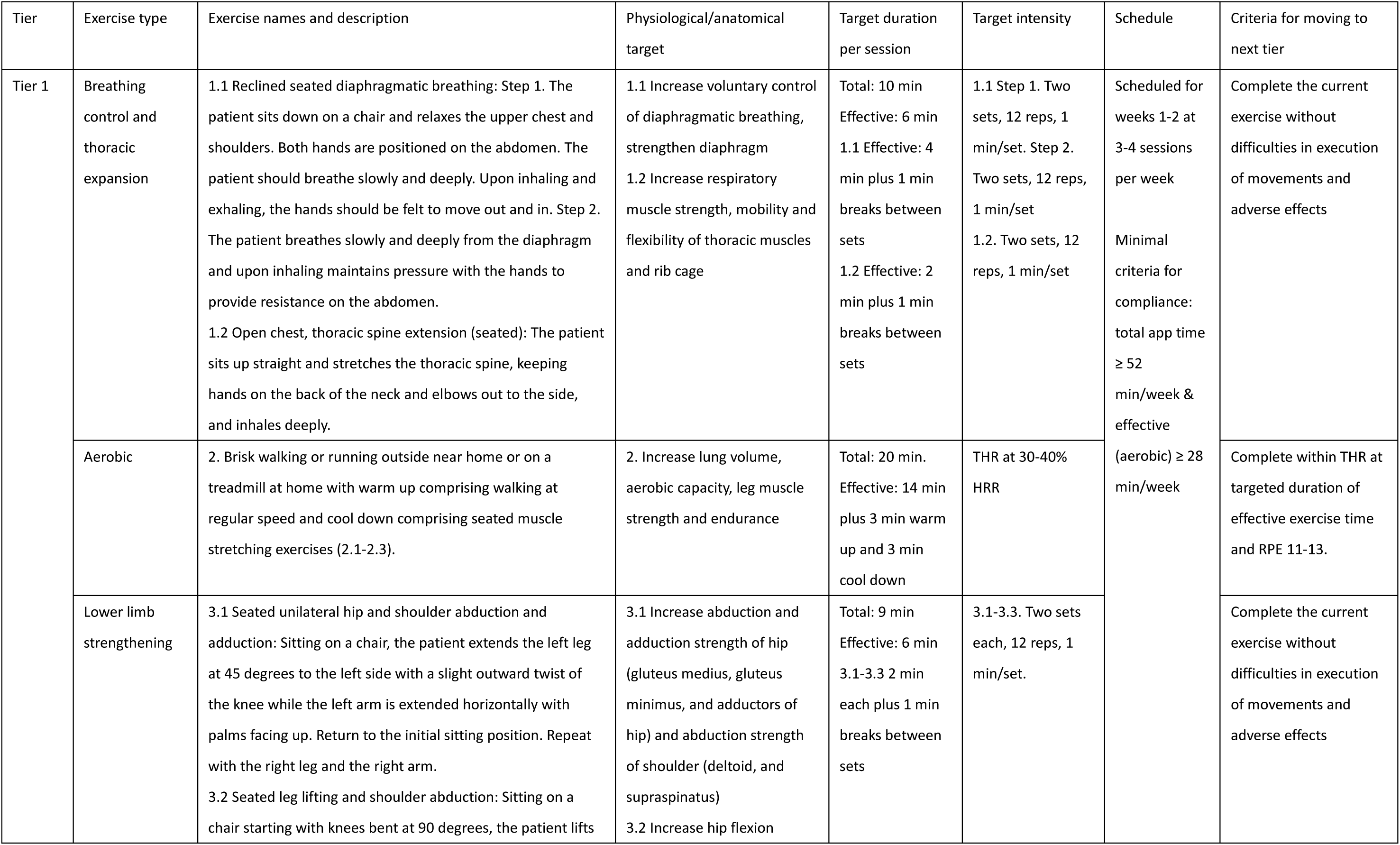
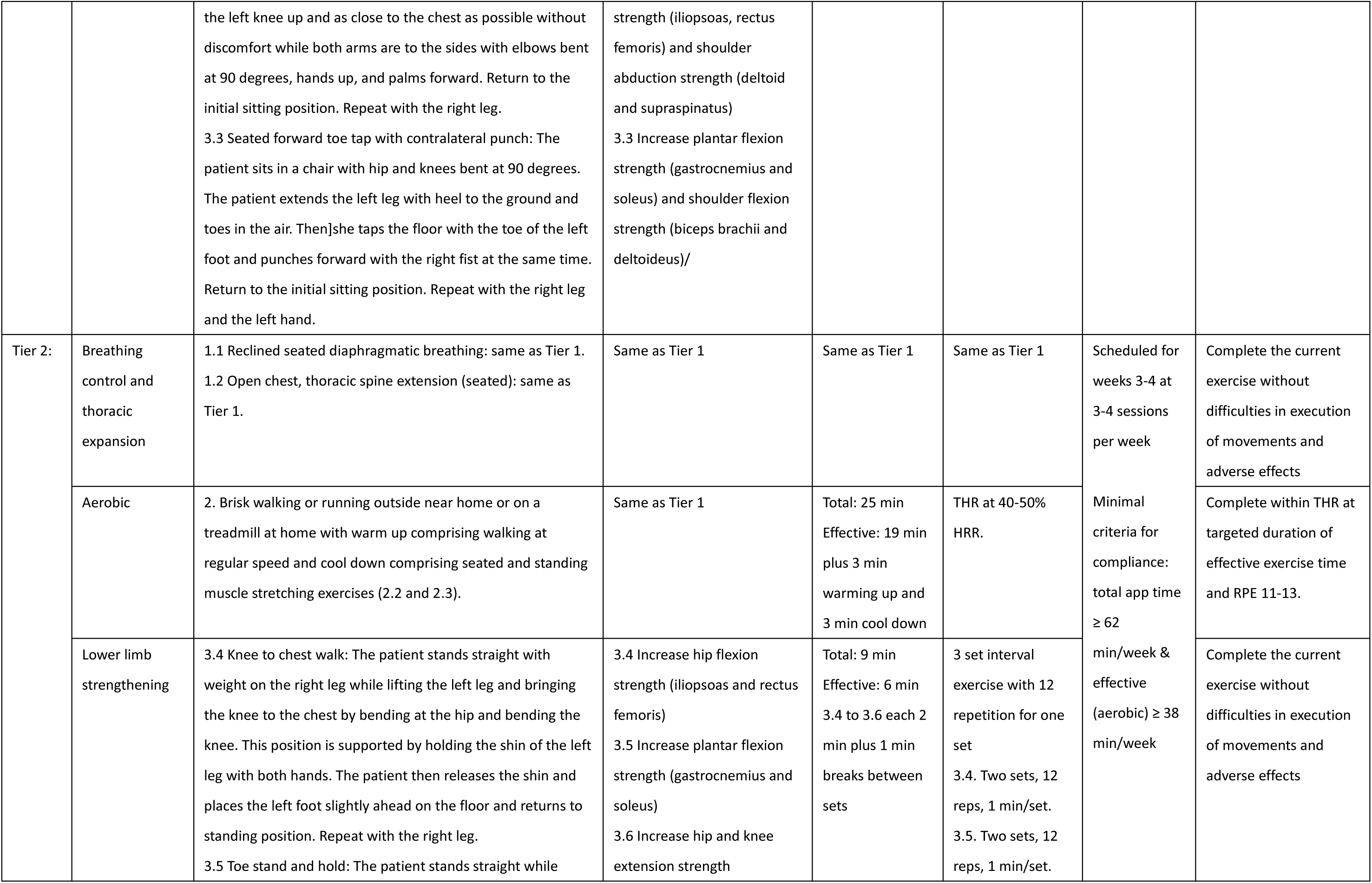
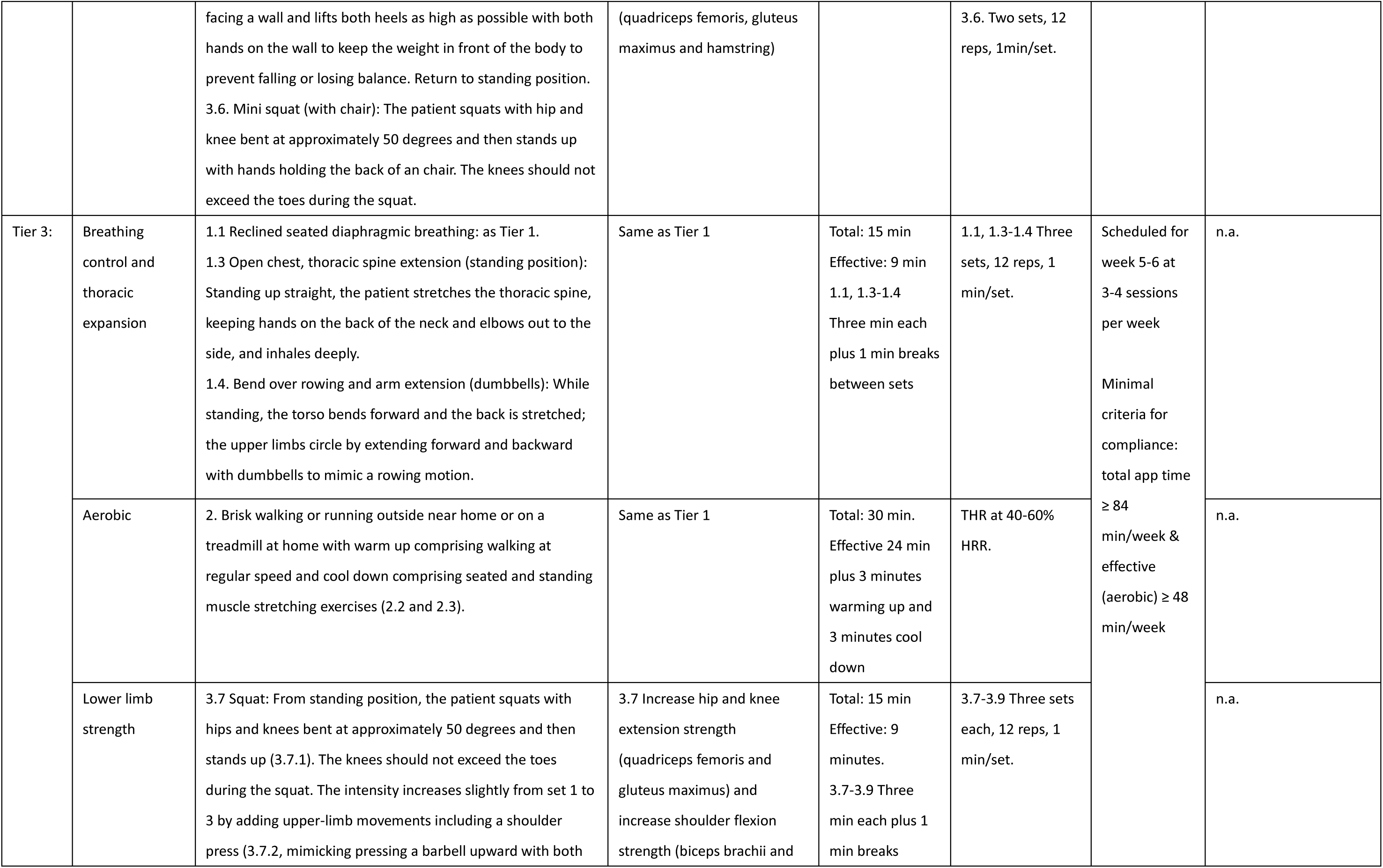
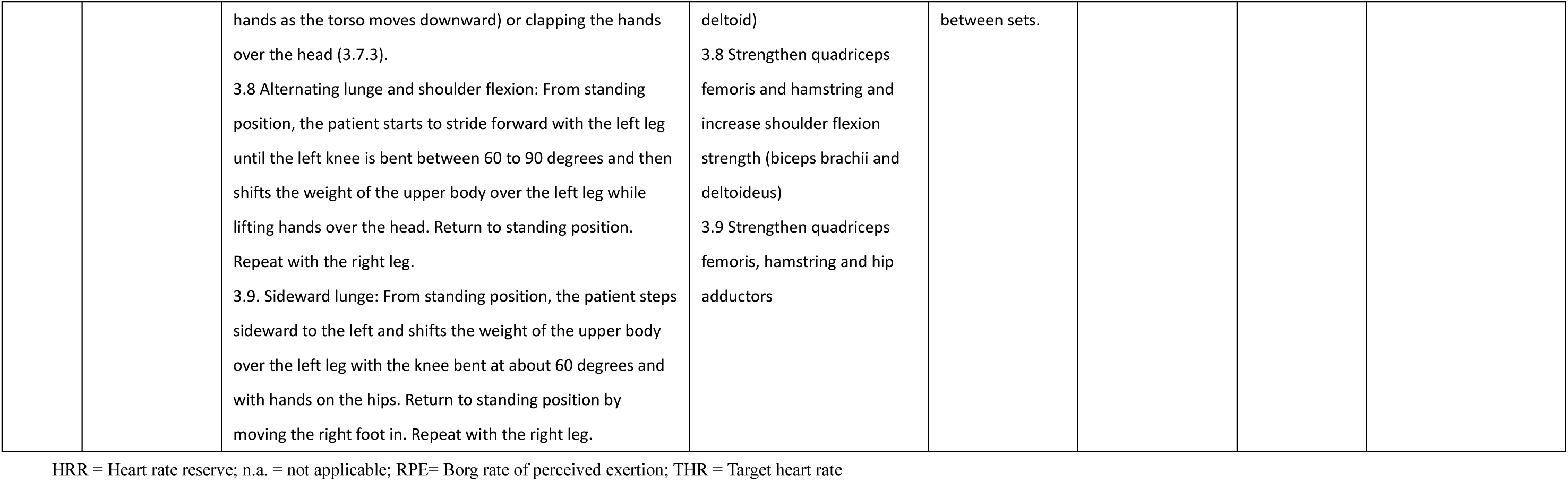
Parameters of three-tiered exercise program for telerehabilitation of Covid-19.

On each day with scheduled exercises, RehabApp reminded the patients to start the exercise and guided them through the process. In the case of BCTE and LMS exercise this was done by displaying videos demonstrating the relevant exercises. During aerobic exercise, information on the patients’ HR in relation to the THR was displayed on the smartphone screen. When the monitored HR was within the THR range, RehabApp recorded the duration as effective exercise and illuminated a green light at the bottom of the screen. When the monitored HR was below or over the THR, a yellow or red light appeared, respectively. If the HR exceeded 110% of the target heart rate, the patient received a message to slow down the exercise until the HR returned to resting rate. RehabApp recorded the HR, the total duration of the exercise, the duration of the effective exercise, self-reported RPE and any interruptions if they occurred.

#### Compliance with exercise protocol

Total exercise time for a given day was determined as the time from opening to closing RehabApp. The time during aerobic exercise within which the monitored HR reached the THR or above was counted as effective exercise time. Having reached at least two thirds (66.7%) of the scheduled total and effective target time as given in Table 1 or modified by the therapist in any given week for at least five of the six weeks according to RehabApp records was considered compliant with the exercise protocol.

### Assessments

Assessments were conducted between 26 April and 9 December 2020. For each patient home visits were scheduled at baseline, at six weeks (post-treatment), and at 24 weeks (follow-up). Additional assessments for perceived dyspnoea and adverse events were performed by consultation via cell phone or WeChat voice call at two weeks and four weeks after baseline. Due to a new policy of the National Health Commission of the PR China announced in the time period between post-treatment and planned follow-up assessments, home visits of patients were no longer permitted for health professionals. Instead, assessments could only be performed after the patients provided negative results of serum antibody and nucleic acid tests for SARS CoV-2. For the final assessment, participants were thus invited to return to the hospitals where they had originally received treatment. Free of charge serum antibody and nucleic acid tests for SARS CoV-2 were provided there. As new appointments for hospital visits had to be scheduled with the patients, this adjustment led to a delay in the final assessment time point for about four weeks on average.

#### Primary outcome

The primary outcome was functional exercise capacity at the post-treatment assessment (6 weeks) measured with the 6-minute walking test (6MWT) administered in accordance with guidelines from the European Respiratory Society and American Thoracic Society^22^ and recorded as 6-minute walking distance (6MWD) in metres. For the first two assessment points a course was arranged outside, near the patients’ home. For the final follow-up assessment a course was arranged in the hospital ward or a hall at the hospital according to the same criteria. If a patient could not complete the 6MWT, the distance walked until interruption was recorded.

#### Secondary outcomes

Secondary outcomes included 6MWD at follow-up, LMS, pulmonary function, health-related quality of life (HRQOL), and perceived dyspnoea. LMS was measured with the squat test^23^. Participants were asked to perform a squat with their back against the wall starting from a standing position, feet flat on the ground, and approximating a 90° angle at the hip and knees. The time in seconds participants could remain in this position was recorded. Pulmonary function was evaluated by spirometry according to guidelines of the American Thoracic Society (Grade C) ^24^. A portable pulmonary function device (MINATO, AS-507, Japan) was used. The following parameters were recorded: forced expiratory volume in the first second (FEV1) in litres, forced vital capacity (FVC) in litres, FEV1/FVC, maximum voluntary ventilation (MVV) in litres per minute, and peak exploratory flow (PEF) in litres per second. For the report of baseline data, the percent of predicted value for FEV1, FVC, and FEV1/FVC, and the percent below lower limit of normal (LLN) were calculated with Global Lung Initiative (GLI-2012) equations for South-East Asia^25^. Percent of predicted PEF and MVV were calculated based on equations for mainland China provided by Mu and Liu^26^. HRQOL was evaluated with the Short Form Health Survey-12 (SF-12) ^27 28^. Physical component score (PCS) and mental component score (MCS) are reported, with higher score indicating better health. Due to the absence of reference equations for mainland China scores were standardised according to US norms^28^. Perceived dyspnoea was assessed with the modified mMRC scale ^29^. Having no symptoms of dyspnoea (mMRC score = 0) was defined as a favourable outcome.

#### Adverse events

Participants could report adverse events including falls, injuries, pain, muscle soreness, fatigue, and other uncomfortable symptoms at any time during the study via phone call or WeChat message to the responsible therapist or researcher. Moreover, participants were asked for any adverse events that had occurred in the previous time period during regular assessments including interim assessments for mMRC dyspnoea at 2 and 4 weeks. Participants in the TERECO group additionally received a prompt by RehabApp after each session asking for adverse events and uncomfortable symptoms that had occurred during exercise. Moreover, they were asked about adverse events in weekly consultations with therapists responsible for monitoring and adapting the intervention. Death, cardiovascular events, other life-threatening events, and re-hospitalisation for events related to the intervention or Covid-19 were defined as a serious adverse event. All reported adverse events were rated by two doctors who had access to the event history and medical history of each patient but were blinded with respect to group allocation. These doctors were not otherwise involved in the study. Severity was rated on a 5-point Likert scale ranging from very mild to life threatening, relationship with the intervention was rated on a 3-point Likert scale (unlikely, possible, likely). Disagreement on particular events was resolved by discussion. If consensus could not be achieved a third reviewer was consulted who had the casting vote.

### Sample size calculation

While a minimal important difference (MID) of 30 metres is recommended for the 6MWT in COPD and other chronic lung disease^30^, an MID for Covid-19 has not yet been established. Given limited knowledge about the longer-term course of the disease including spontaneous recovery as well as learning effects that may occur, a more conservative MID of 50 metres was assumed here. To achieve 80% power with an alpha error of 5%, a minimal sample size of 96 participants (48 per group) was needed to detect a statistically significant signal for a between-group difference in change in 6MWD (with SD 99 in control and SD 71 in the intervention group as reported by Lau for a trial in a SARS population^11^). Assuming 20% attrition our recruitment target was 120 participants (60 per group).

### Statistical analysis

All analyses were performed with STATA 14.0. All main analyses were conducted on intention-to-treat (ITT) basis and without imputations. Statistical significance was set at alpha = 5% with two-sided tests. No corrections for multiplicity were applied. The predefined statistical analysis plan (SAP) is available in Appendix 1.

#### Main analysis

Data were assumed to be missing at random (MAR) with missing values depending on observed model parameters^31^. This assumption was tested with sensitivity analysis as described below. The primary outcome, change in 6MWD post-treatment, was evaluated with constrained longitudinal data analysis (cLDA), that is a linear mixed effects model that imposed an equality constraint on baseline means^32 33^. Dependence of longitudinal observations within study participants was modelled with a random intercept. The fixed part of the model included time points and interactions between time points and treatment group (but no term for treatment group at baseline), and a term for study centre. This model is equivalent to longitudinal analysis of covariance (ANCOVA) if there are no missing data. However, in the presence of missing data, cLDA also considers data from participants who only participated in the baseline assessment. The parameters for the interaction terms between time and treatment represent the estimated treatment effects at the two follow-up points and are reported with 95% confidence intervals (CIs). In contrast to the SAP, treatment effects for post-treatment and follow-up assessment were estimated simultaneously, within the same model. This was done because two participants who had missed the post-treatment assessment returned for the follow-up assessment and using this information for model estimation was considered important in light of the principles of ITT analysis.

All secondary outcomes apart from mMRC dyspnoea were analysed analogously. Occurrence of a favourable outcome in mMRC dyspnoea was analysed with a generalised linear model of the Poisson family with a log link. This model used the natural logarithm of the number of observed occasions until the respective data point as offset and was adjusted for centre (fixed effect). Cluster robust standard errors (cluster variable: participant ID) were used for the estimation of 95% CIs^34^. Baseline was not modelled for mMRC dyspnoea since all participants did have dyspnoea at baseline (see inclusion criteria). Aside from terms for post-treatment and follow-up assessment, and their interaction with treatment group, two additional time points (for weeks 2 and 4) and their interactions with treatment were included. Treatment effects are presented as rate ratios (RR) along with logistic 95% CIs. The Poisson model was chosen over a longitudinal logistic model originally specified in the SAP due to ease of interpretation of rate ratios and derivation of population-averaged probabilities. A graphical illustration of trajectories presents marginal means and probabilities (mMRC dyspnoea) for the intervention group and time point as estimated with the above models with 84% CIs serving as comparisons bars. 84% CIs allow for visual inspection for approximate statistical significance of mean differences at alpha = 5% by looking at the overlap of CIs.^35^

#### Sensitivity analysis

Pre-specified sensitivity analysis included estimation of the above models on the per protocol sample, as well as on two types of multiply imputed datasets. Firstly, multiple imputation with chained equations^36^ was performed under an extended MAR assumption, i.e. that missing values were also dependent on observed values of (auxiliary) variables not included in the models used for primary analysis. These were gender, age, disease severity, time from first hospital admission for Covid-19 to baseline assessment, presence of comorbidities, smoking history, and body mass index. Models were estimated on 70 imputed data sets as the originally-specified 50 data sets (see SAP) did not yield a satisfactory upper limit for the fraction of missing information^37^. Secondly, controlled multiple imputation (50 sets) was used for simulating a non-MAR scenario where patients with missing assessments in the TERECO group followed the pattern of change in controls (copy increments in reference - CIR).^38^

As the follow-up assessment could not take place at home as planned due to change in administrative regulations pertaining to Covid-19 as previously noted, a delay in the planned follow-up assessment occurred for several patients and unequal time periods between post-treatment and follow-up assessment resulted. Moreover, there was considerable variation in days from onset of symptoms to admission (mean 7.4, SD 9.8, range 0-70) which was unexpected. For these reasons, additional post hoc sensitivity analysis was conducted; the models outlined under main analysis were fitted with two additional terms for time since onset of symptoms (TOS) and TOS squared.

#### Analysis of harms

Adverse events were descriptively analysed.

### Summary of protocol deviations

(i) Follow-up assessments could not be conducted by home visit due to change in administrative regulations regarding Covid-19, but had to be performed at the participating hospitals. Consequently, follow-up assessments could not be completed at 24 weeks as planned but were conducted at 28 weeks after baseline on average. (ii) The primary outcome 6MWD at post-treatment assessment was analysed simultaneously with the secondary outcome 6MWD at follow-up, i.e. a joint model was fit as opposed to two separate models. (iii) mMRC perceived dyspnoea (favourable outcome) was analysed with log-linear Poisson regression with cluster-robust standard errors instead of longitudinal logistic regression. (iv) Multiple imputations for sensitivity analysis (extended MAR scenario) were performed on 70 data sets and not 50 sets. Deviations from the SAP (ii-iv) had no effect on the statistical significance of estimates when compared with analyses as originally planned.

### Patient involvement

Patients were not systematically involved in the design of this study. However, conversations with patients and their families concerning what goals they had after discharge and what kind of exercise they could perform at home informed the design of the TERECO program. Moreover, extensive feedback from individual patients allocated to the TERECO group was collected during the intervention period through RehabApp and in weekly consultations with therapists to better adapt individual exercises to patients’ needs and reduce any discomfort during and after training sessions. Results will be disseminated to patients in both study groups upon publication.

## RESULTS

**Figure 1** illustrates the flow of patients through the study. After pre-screening of hospital records, 140 patients were contacted for further evaluation of eligibility between 22 April and May 28 2020. Of those 20 were ineligible or refused consent. 120 patients were randomised with one person in the TERECO group and one in the control group not receiving the allocated intervention. One patient was withdrawn from the TERECO group before the start of the exercise program as a premature beat was discovered. One patient in the control group had been randomised mistakenly because the assessor forgot to inform the allocator about the patient’s ineligibility due to refusal to collaborate in the baseline 6MWT and pulmonary function assessment (see exclusion criterion vi). Other data from this patient were not included in the analysis. Six patients of the TERECO group did not complete the post-treatment assessment. We lost contact with four patients of which one had previously announced to discontinue the intervention for unspecified reasons. Two patients who discontinued the intervention, one because of chest pain and one for unspecified reasons, missed the post-treatment assessment but returned for the follow-up assessment. We had lost contact with four additional patients in the TERECO group and five patients from the control group at the final follow-up point.

Baseline characteristics of the study participants by intervention group are provided in Table 2. The overall mean age of the study population was 50.61 (SD 10.98), 53 (44.5%) were male, and 73 (61.3%) had at least one comorbidity. Length of hospital stay for acute treatment was 26.18 on average (SD 15.25) with a mean of 70 days (SD 16.85) from hospital discharge to baseline. Fifty (43.5%) patients were below LLN for FEV1, 45 (39.1%) for FVC, and 26 (22.6%) for FEV1/FVC.

**Table 2:**
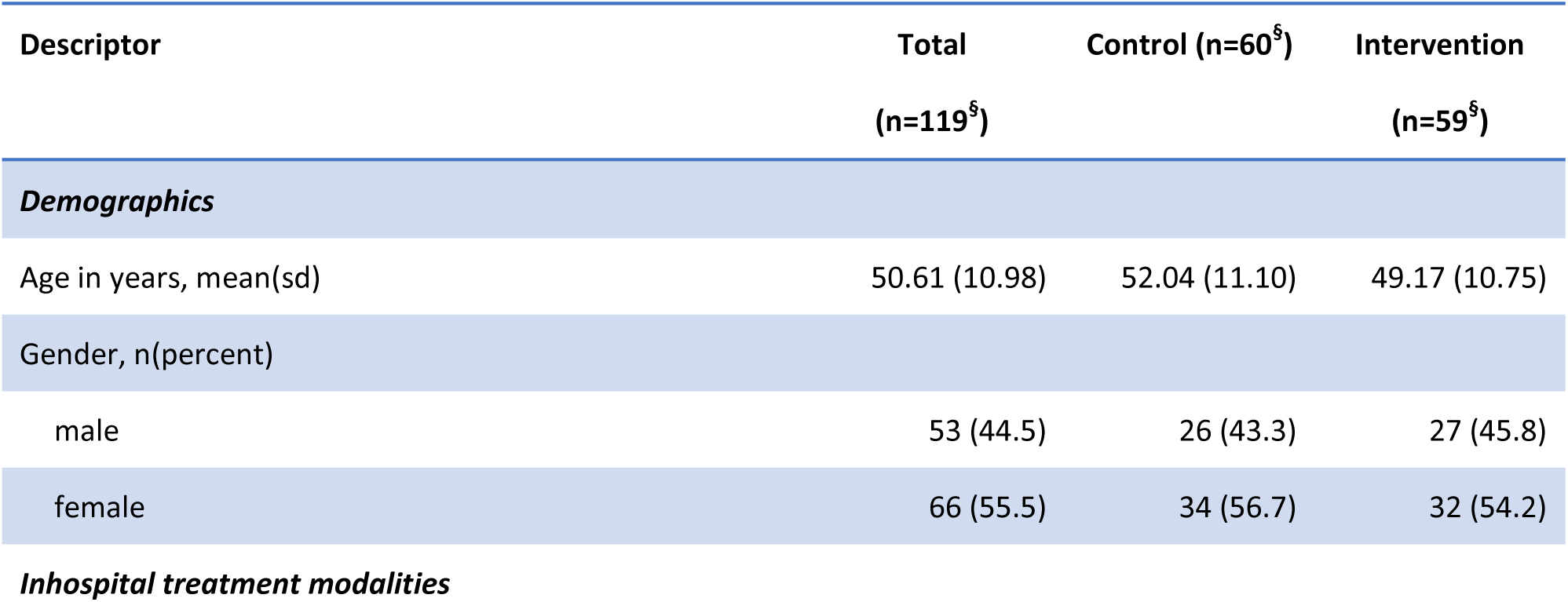
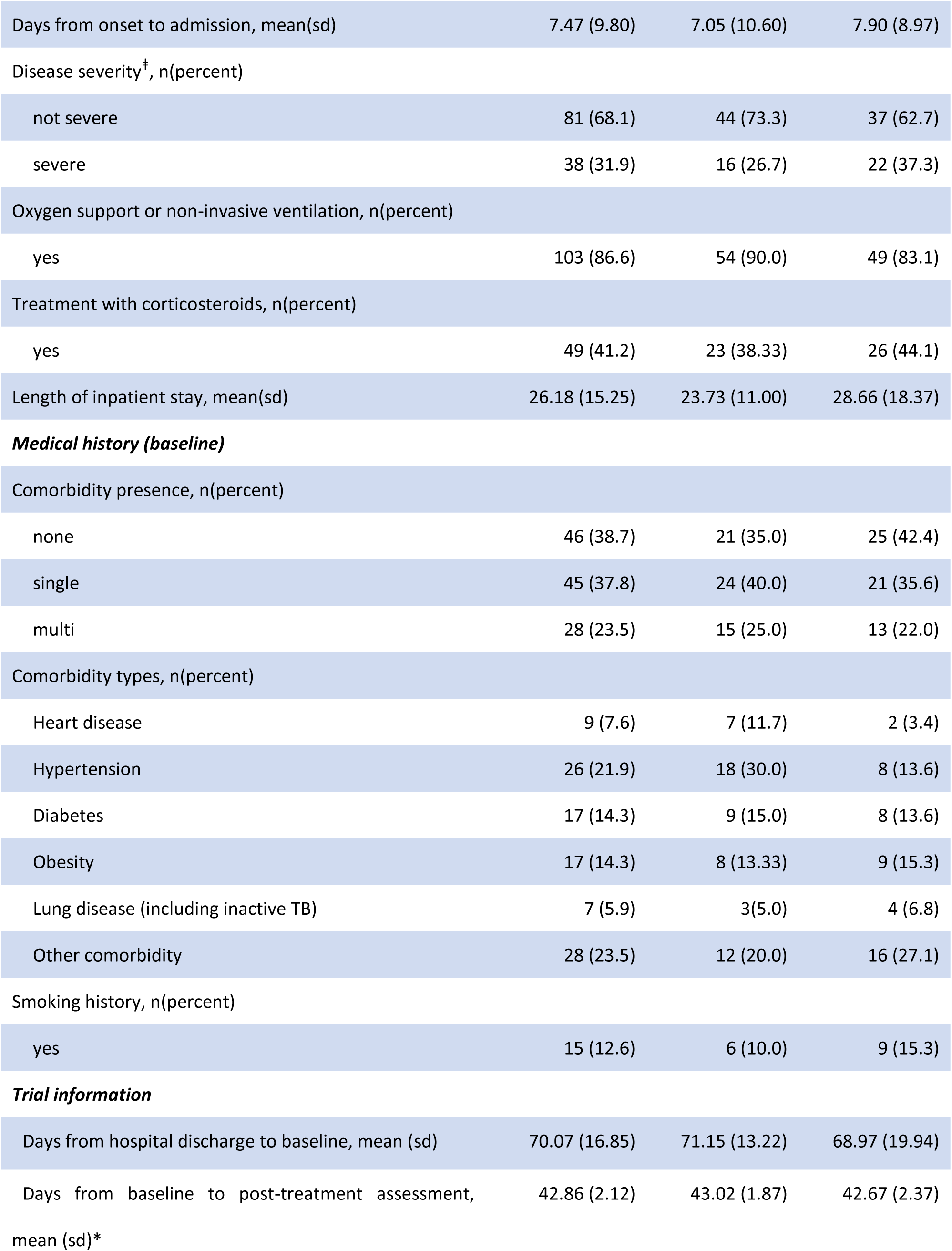
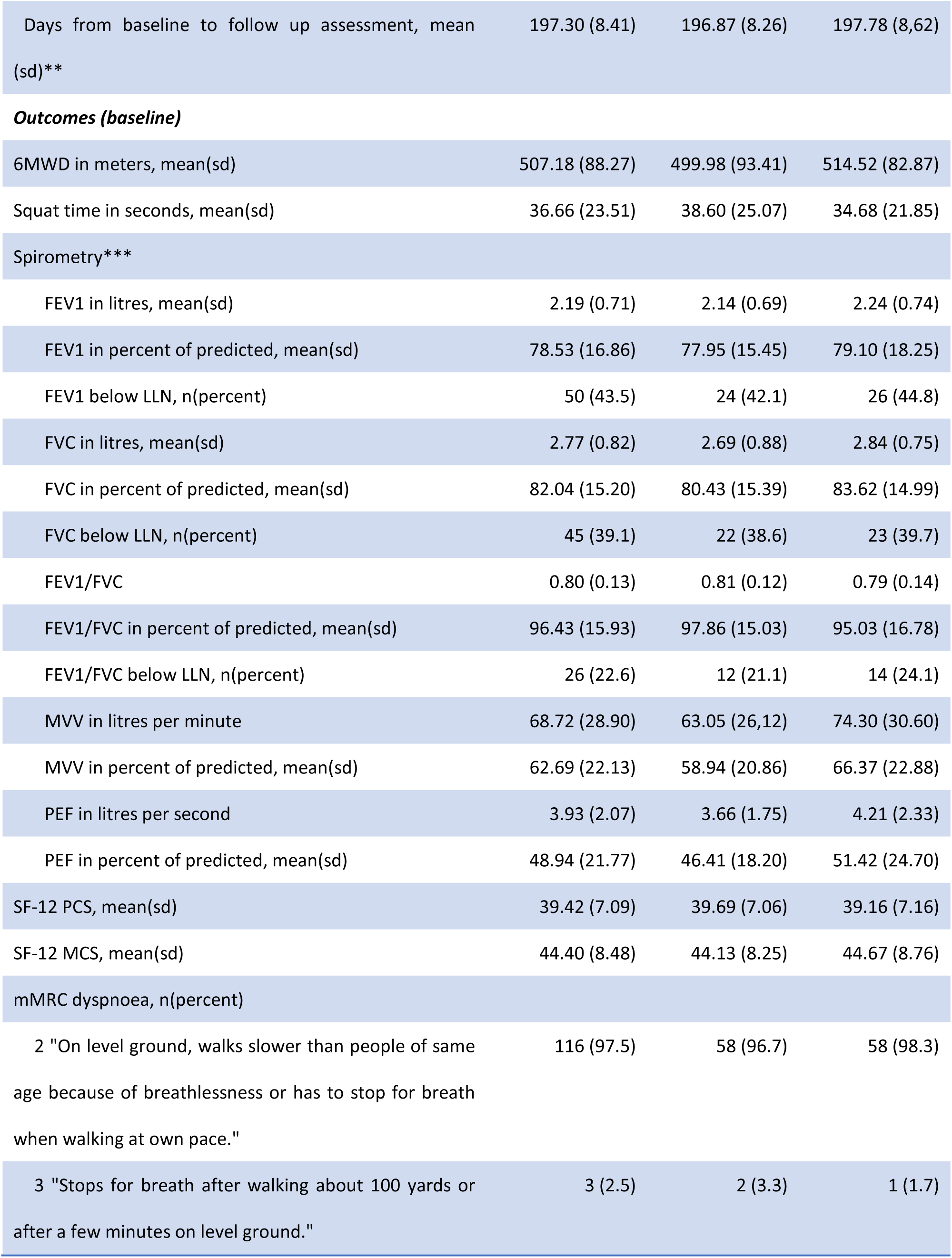
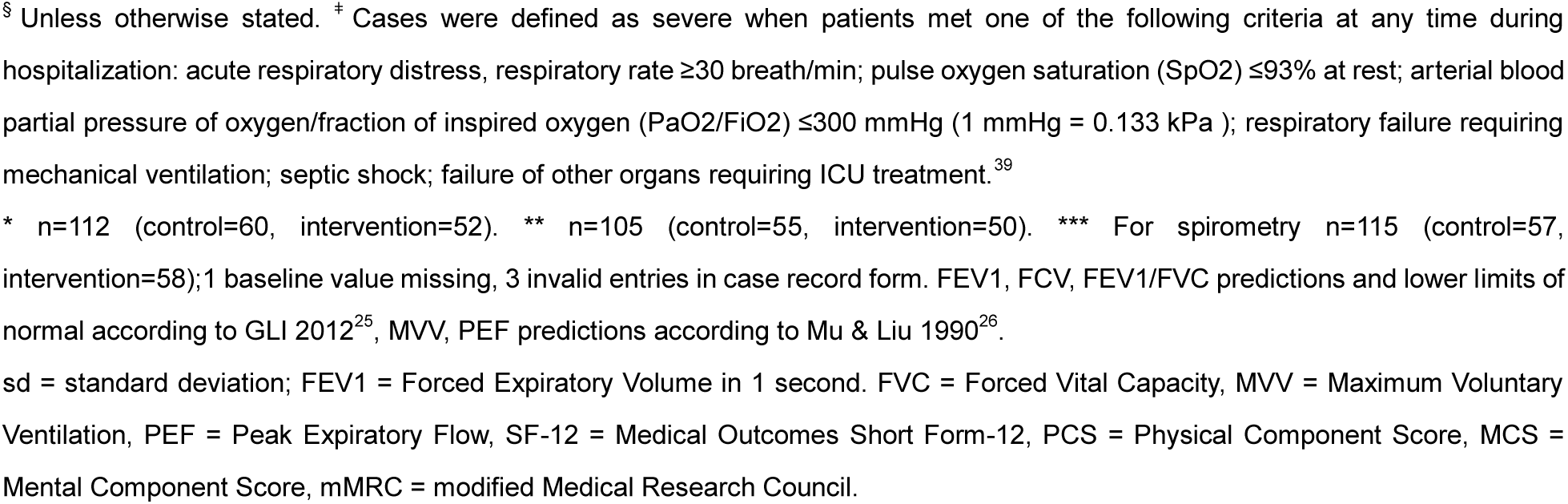
Participant characteristics and outcomes at baseline.

| Descriptor | Total<br>(n=119<sup>§</sup>) | Control (n=60<sup>§</sup>) | Intervention<br>(n=59<sup>§</sup>) |
| --- | --- | --- | --- |
| **Demographics** |  |  |  |
| Age in years, mean(sd) | 50.61 (10.98) | 52.04 (11.10) | 49.17 (10.75) |
| Gender, n(percent) |  |  |  |
| male | 53 (44.5) | 26 (43.3) | 27 (45.8) |
| female | 66 (55.5) | 34 (56.7) | 32 (54.2) |
| **Inhospital treatment modalities** |  |  |  |
| Days from onset to admission, mean(sd) | 7.47 (9.80) | 7.05 (10.60) | 7.90 (8.97) |
| Disease severity<sup>†</sup>, n(percent) |  |  |  |
| not severe | 81 (68.1) | 44 (73.3) | 37 (62.7) |
| severe | 38 (31.9) | 16 (26.7) | 22 (37.3) |
| Oxygen support or non-invasive ventilation, n(percent) |  |  |  |
| yes | 103 (86.6) | 54 (90.0) | 49 (83.1) |
| Treatment with corticosteroids, n(percent) |  |  |  |
| yes | 49 (41.2) | 23 (38.33) | 26 (44.1) |
| Length of inpatient stay, mean(sd) | 26.18 (15.25) | 23.73 (11.00) | 28.66 (18.37) |
| **Medical history (baseline)** |  |  |  |
| Comorbidity presence, n(percent) |  |  |  |
| none | 46 (38.7) | 21 (35.0) | 25 (42.4) |
| single | 45 (37.8) | 24 (40.0) | 21 (35.6) |
| multi | 28 (23.5) | 15 (25.0) | 13 (22.0) |
| Comorbidity types, n(percent) |  |  |  |
| Heart disease | 9 (7.6) | 7 (11.7) | 2 (3.4) |
| Hypertension | 26 (21.9) | 18 (30.0) | 8 (13.6) |
| Diabetes | 17 (14.3) | 9 (15.0) | 8 (13.6) |
| Obesity | 17 (14.3) | 8 (13.33) | 9 (15.3) |
| Lung disease (including inactive TB) | 7 (5.9) | 3(5.0) | 4 (6.8) |
| Other comorbidity | 28 (23.5) | 12 (20.0) | 16 (27.1) |
| Smoking history, n(percent) |  |  |  |
| yes | 15 (12.6) | 6 (10.0) | 9 (15.3) |
| **Trial information** |  |  |  |
| Days from hospital discharge to baseline, mean (sd) | 70.07 (16.85) | 71.15 (13.22) | 68.97 (19.94) |
| Days from baseline to post-treatment assessment, mean (sd)* | 42.86 (2.12) | 43.02 (1.87) | 42.67 (2.37) |
| Days from baseline to follow up assessment, mean (sd)** | 197.30 (8.41) | 196.87 (8.26) | 197.78 (8.62) |
| **Outcomes (baseline)** |  |  |  |
| 6MWD in meters, mean(sd) | 507.18 (88.27) | 499.98 (93.41) | 514.52 (82.87) |
| Squat time in seconds, mean(sd) | 36.66 (23.51) | 38.60 (25.07) | 34.68 (21.85) |
| **Spirometry\*\*\*** |  |  |  |
| FEV1 in litres, mean(sd) | 2.19 (0.71) | 2.14 (0.69) | 2.24 (0.74) |
| FEV1 in percent of predicted, mean(sd) | 78.53 (16.86) | 77.95 (15.45) | 79.10 (18.25) |
| FEV1 below LLN, n(percent) | 50 (43.5) | 24 (42.1) | 26 (44.8) |
| FVC in litres, mean(sd) | 2.77 (0.82) | 2.69 (0.88) | 2.84 (0.75) |
| FVC in percent of predicted, mean(sd) | 82.04 (15.20) | 80.43 (15.39) | 83.62 (14.99) |
| FVC below LLN, n(percent) | 45 (39.1) | 22 (38.6) | 23 (39.7) |
| FEV1/FVC | 0.80 (0.13) | 0.81 (0.12) | 0.79 (0.14) |
| FEV1/FVC in percent of predicted, mean(sd) | 96.43 (15.93) | 97.86 (15.03) | 95.03 (16.78) |
| FEV1/FVC below LLN, n(percent) | 26 (22.6) | 12 (21.1) | 14 (24.1) |
| MVV in litres per minute | 68.72 (28.90) | 63.05 (26.12) | 74.30 (30.60) |
| MVV in percent of predicted, mean(sd) | 62.69 (22.13) | 58.94 (20.86) | 66.37 (22.88) |
| PEF in litres per second | 3.93 (2.07) | 3.66 (1.75) | 4.21 (2.33) |
| PEF in percent of predicted, mean(sd) | 48.94 (21.77) | 46.41 (18.20) | 51.42 (24.70) |
| SF-12 PCS, mean(sd) | 39.42 (7.09) | 39.69 (7.06) | 39.16 (7.16) |
| SF-12 MCS, mean(sd) | 44.40 (8.48) | 44.13 (8.25) | 44.67 (8.76) |
| **mMRC dyspnoea, n(percent)** |  |  |  |
| 2 "On level ground, walks slower than people of same age because of breathlessness or has to stop for breath when walking at own pace." | 116 (97.5) | 58 (96.7) | 58 (98.3) |
| 3 "Stops for breath after walking about 100 yards or after a few minutes on level ground." | 3 (2.5) | 2 (3.3) | 1 (1.7) |

§ Unless otherwise stated. \* Cases were defined as severe when patients met one of the following criteria at any time during hospitalization: acute respiratory distress, respiratory rate $\geq 30$ breath/min; pulse oxygen saturation (SpO<sub>2</sub>) $\leq 93\%$ at rest; arterial blood partial pressure of oxygen/fraction of inspired oxygen (PaO<sub>2</sub>/FiO<sub>2</sub>) $\leq 300$ mmHg (1 mmHg = 0.133 kPa); respiratory failure requiring mechanical ventilation; septic shock; failure of other organs requiring ICU treatment.<sup>39</sup>
\* n=112 (control=60, intervention=52). \*\* n=105 (control=55, intervention=50). \*\*\* For spirometry n=115 (control=57, intervention=58); 1 baseline value missing, 3 invalid entries in case record form. FEV<sub>1</sub>, FVC, FEV<sub>1</sub>/FVC predictions and lower limits of normal according to GLI 2012<sup>25</sup>, MVV, PEF predictions according to Mu & Liu 1990<sup>26</sup>.
sd = standard deviation; FEV<sub>1</sub> = Forced Expiratory Volume in 1 second. FVC = Forced Vital Capacity, MVV = Maximum Voluntary Ventilation, PEF = Peak Expiratory Flow, SF-12 = Medical Outcomes Short Form-12, PCS = Physical Component Score, MCS = Mental Component Score, mMRC = modified Medical Research Council.

Overall, 38 participants in the TERECO group complied with the exercise protocol, 64.41% of those randomized and 73.3% of those who remained in the program for the full six weeks.

### Primary outcome

The mean 6MWD in the control group improved by 17.1 metres (SD 63.9) from baseline to post-treatment assessment at 6 weeks, whereas 6MWD in the TERECO group increased by 80.2 metres (SD 74.7). The adjusted between-group difference in change in 6MWD from baseline (treatment effect) was 65.45 metres (95% CI 43.8-87.1, p<0.001).

### Secondary outcomes

Table 3 gives an overview of crude change and adjusted treatment effects for all outcomes.

**Table 3:** Crude change in outcomes from baseline at the different assessment points and estimated adjusted treatment effects with 95% confidence intervals (on intention to treat basis)

|  | Number of participants |  | Crude change from baseline<sup>§</sup> |  | Estimated treatment effect<sup>‡</sup> (95% CI) | P |
| --- | --- | --- | --- | --- | --- | --- |
|  | Control | TERECO | Control | TERECO |  |  |
| Primary outcome |  |  |  |  |  |  |
| 6MWD in meters |  |  |  |  |  |  |
| - post-treatment (6 weeks) | 60 | 52 | 17.09 ± 63.94 | 80.20 ± 74.66 | 65.45 (43.80-87.10) | < 0.001 |
| - follow up (~28 weeks) | 55 | 50 | 15.17 ± 70.02 | 84.81 ± 80.38 | 68.62 (46.39-90.85) | < 0.001 |
| Secondary outcomes |  |  |  |  |  |  |
| Squat time in seconds |  |  |  |  |  |  |
| - post-treatment (6 weeks) | 60 | 52 | 7.98 ± 19.53 | 29.35 ± 27.22 | 20.12 (12.34-27.90) | < 0.001 |
| - follow up (~28 weeks) | 55 | 50 | 4.16 ± 19.62 | 28.12 ± 27.17 | 22.23 (14.24-30.21) | < 0.001 |
| Pulmonary function |  |  |  |  |  |  |
| FEV1 in litres |  |  |  |  |  |  |
| - post-treatment (6 weeks) | 56 | 51 | 0.18 ± 0.53 | 0.28 ± 0.51 | 0.08 (-0.08-0.25) | 0.327 |
| - follow up (~28 weeks) | 53 | 47 | 0.29 ± 0.43 | 0.29 ± 0.48 | 0.00 (-0.18-0.17) | 0.969 |
| FVC in litres |  |  |  |  |  |  |
| - post-treatment (6 weeks) | 56 | 51 | 0.19 ± 0.40 | 0.21 ± 0.47 | 0.02 (-0.14-0.18) | 0.818 |
| - follow up (~28 weeks) | 53 | 47 | 0.27 ± 0.43 | 0.30 ± 0.38 | 0.01 (-0.16-0.17) | 0.95 |
| FEV1/FVC |  |  |  |  |  |  |
| - post-treatment (6 weeks) | 56 | 51 | 0.01 ± 0.16 | 0.04 ± 0.17 | 0.03 (-0.02-0.07) | 0.224 |
| - follow up (~28 weeks) | 53 | 47 | 0.02 ± 0.15 | 0.02 ± 0.18 | -0.01 (-0.05-0.03) | 0.732 |
| MVV in litres per minute |  |  |  |  |  |  |
| - post-treatment (6 weeks) | 56 | 51 | 5.61 ± 17.31 | 14.49 ± 21.60 | 10.57 (3.26-17.88) | 0.005 |
| - follow up (~28 weeks) | 53 | 47 | 13.81 ± 20.78 | 18.47 ± 22.31 | 5.20 (-2.33-12.73) | 0.176 |
| PEF in litres per second |  |  |  |  |  |  |
| - post-treatment (6 weeks) | 56 | 51 | 0.66 ± 1.95 | 0.98 ± 1.90 | 0.38 (-0.24-1.00) | 0.229 |
| - follow up (~28 weeks) | 53 | 47 | 0.97 ± 1.84 | 0.76 ± 1.92 | -0.02 (-0.66-0.62) | 0.954 |
| *Quality of life* |  |  |  |  |  |  |
| SF-12 PCS |  |  |  |  |  |  |
| - post-treatment (6 weeks) | 60 | 52 | 3.84 ± 7.60 | 7.81 ± 7.02 | 3.79 (1.24-6.35) | 0.004 |
| - follow up (~28 weeks) | 55 | 50 | 5.20 ± 9.13 | 8.2 ± 10.05 | 2.69 (0.06-5.32) | 0.045 |
| SF-12 MCS |  |  |  |  |  |  |
| - post-treatment (6 weeks) | 60 | 52 | 4.17 ± 8.79 | 6.15 ± 10.78 | 2.18 (-0.54-4.90) | 0.116 |
| - follow up (~28 weeks) | 55 | 50 | 5.51 ± 7.79 | 6.92 ± 10.28 | 1.99 (-0.81-4.79) | 0.164 |
| *mMRC perceived dyspnoea, favourable outcome* |  |  |  |  |  |  |
| - interim 2 weeks | 60 | 54 | 45.0 | 57.4 | 1.27 (0.88-1.82) | 0.197 |
| - interim 4 weeks | 60 | 54 | 61.7 | 66.7 | 1.08 (0.82-1.42) | 0.605 |
| - post-treatment (6 weeks) | 60 | 52 | 61.7 | 90.4 | 1.46 (1.17-1.82) | 0.001 |
| - follow up (~28 weeks) | 55 | 50 | 60.0 | 72.0 | 1.22 (0.92-1.61) | 0.162 |

§Crude change from baseline is given as mean change ± standard deviation of this change for all outcomes apart from mMRC (favorable outcome); for the latter outcome percentages endorsing a score of zero (no dyspnea) at each time point are provided.
‡Estimated treatment effects for all outcomes apart from mMRC dyspnoea (favorable outcome) are between-group mean differences in change from baseline derived from mixed effects regression with random intercept for study participant; models are constrained to a common baseline mean across groups and adjusted for centre. Estimation includes all available observations from participants randomized (number of participants with valid observations at baseline is 115 for pulmonary function parameters and 119 for all other outcomes). Estimated treatment effects for mMRC dyspnoea (favourable outcome) are risk ratios derived from generalized linear model from Poisson family with log link adjusted for centre and ln(number valid observations up to data point) as offset; 95% CIs are based on cluster robust standard errors (cluster variable: participant ID).
TERECO = telerehabilitation intervention for Covid-19 survivors, CI = confidence interval, 6MWD = six minute walking distance, FEV1 = forced expiratory volume in 1 second, FVC = forced vital capacity, MVV = maximum voluntary ventilation, PEF = peak expiratory flow, SF-12 = Medical Outcomes Short Form-12, PCS = physical component score, MCS = mental component score, mMRC = modified Medical Research Council

**Figure 2** depicts marginal trajectories over time by study group with 84% CIs (comparison bars) serving as a visual aid for inspecting the statistical significance of mean differences at the 5% level (see Appendix 2 for detailed estimates).

With estimated 68.62 metres (95% CI 46.39-90.85, p<0.001) the treatment effect regarding 6MWD increased somewhat at follow-up. LMS improved to a larger degree in the TERECO group as compared to control with estimated treatment effects of 20.12 seconds in squat position (95% CI 12.34-27.9, p<0.001) post-treatment, and 22.23 seconds (95% CI 14.24-30.21, p<0.001) at follow-up. Lung function parameters improved in both group over time (Figure 2). No group differences were found apart from an adjusted between-group difference in change from baseline of 10.57 litres per minute (95% CI 3.26-17.88, p=0.005) in post-treatment MVV in favour of the TERECO group. This between-group difference was reduced to 5.2 litres per minute (95% CI −2.33-12.73) at follow-up and was no longer statistically significant (p=0.176). SF-12 PCS increased to a larger degree in the TERECO group as compared to the control group with treatment effects estimated as 3.79 points (95% CI 1.24-6.35, p=0.004) at the post-treatment assessment and 2.69 (95% CI 0.06-5.32, p=0.045) at follow-up. The lower limit of the 95% CI of the between-group difference was however close to zero at follow-up. Improvement in SF-12 MCS was somewhat greater in the TERECO group but 95% CIs were also compatible with greater improvement in the control group at both assessment points. With 90.4% endorsing a favourable outcome for dyspnoea in the TERECO group as opposed to 61.7 percent in the control group (adjusted RR 1.46, 1.17-1.82, p=0.001), a treatment effect for perceived dyspnoea was found immediately after the intervention period but not at the other time points.

### Sensitivity analysis

Estimates from sensitivity analysis are provided in Table 4. Per protocol analysis showed larger effect estimates for 6MWD and LMS. For the per protocol sample the estimated between-group difference in change from baseline for the primary outcome was 72.25 metres (95% CI 47.54-96.97, p>0.001). With the exception of 6MWD at follow-up, treatment effects were lowest under the CIR scenario, followed by the extended MAR scenario. The lowest treatment effect for the primary outcome was 57.18 metres (35.42-78.95, p<0.001) under the CIR scenario. The extended MAR scenario showed the lowest adjusted treatment effect for 6MWD at follow-up with 61.99 metres (95% CI 39.22-84.76, p<0.001). A longer-term effect of TERECO on SF-12 PCS was unstable in all pre-planned sensitivity analyses. Estimates from post hoc sensitivity analysis that added parameters for TOS to the models were almost identical with those from main analysis.

**Table 4:** Results of sensitivity analysis.

| Outcome | Estimates of treatment effects from different scenarios with 95% CIs |  |  |  |  |
| --- | --- | --- | --- | --- | --- |
|  | ITT, primary analysis<br>(n=119, n<sub>int</sub>=59, n<sub>obs</sub> = 336) | Per protocol*<br>(n=91, n<sub>int</sub>=36) | ITT, extended MAR<br>multiple imputation<sup>§</sup><br>(n=119, n<sub>int</sub>=59, 70 sets) | ITT, CIR multiple<br>imputation<sup>‡</sup><br>(n=119, n<sub>int</sub>=59, 50 sets) | ITT, model including<br>time from symptoms<br>onset<sup>&</sup><br>(n=119, n<sub>int</sub>=59, n<sub>obs</sub> = 336) |
| **Primary outcome: 6MWD in meters** |  |  |  |  |  |
| - post-treatment (6 weeks) | 65.45 (43.80-87.10) | 72.25 (47.54-96.97) | 62.23 (40.07-84.39) | 57.18 (35.42-78.95) | 65.12 (44.50-85.74) |
| - follow up (~28 weeks) | 68.62 (46.39-90.85) | 75.92 (51.21-100.64) | 61.99 (39.22-84.76) | 63.07 (40.87-85.27) | 67.99 (46.77-89.20) |
| **Secondary outcomes** |  |  |  |  |  |
| *Squat time in seconds* |  |  |  |  |  |
| - post-treatment (6 weeks) | 20.12 (12.34-27.90) | 22.67 (13.91-31.43) | 20.32 (11.72-28.91) | 17.81 (10.01-25.61) | 20.15 (12.44-27.86) |
| - follow up (~28 weeks) | 22.23 (14.24-30.21) | 25.94 (17.18-34.70) | 21.48 (12.73-30.24) | 20.07 (12.09-28.06) | 22.38 (14.45-30.31) |
| *Pulmonary function* |  |  |  |  |  |
| FEV1 in litres |  |  |  |  |  |
| - post-treatment (6 weeks) | 0.08 (-0.08-0.25) | 0.09 (-0.1-0.28) | 0.07 (-0.11-0.25) | 0.06 (-0.11-0.24) | 0.08 (-0.08-0.25) |
| - follow up (~28 weeks) | 0.00 (-0.18-0.17) | -0.03 (-0.22-0.16) | -0.05 (-0.23-0.13) | 0.00 (-0.18-0.18) | 0.00 (-0.18-0.17) |
| FVC in litres |  |  |  |  |  |
| - post-treatment (6 weeks) | 0.02 (-0.14-0.18) | 0.09 (-0.08-0.27) | -0.01 (-0.18-0.16) | 0.03 (-0.13-0.20) | 0.02 (-0.14-0.18) |
| - follow up (~28 weeks) | 0.01 (-0.16-0.17) | 0.07 (-0.11-0.25) | -0.06 (-0.23-0.12) | 0.01 (-0.16-0.18) | 0.01 (-0.15-0.17) |
| FEV1/FVC |  |  |  |  |  |
| - post-treatment (6 weeks) | 0.03 (-0.02-0.07) | 0.02 (-0.02-0.07) | 0.02 (-0.02-0.06) | 0.02 (-0.02-0.06) | 0.02 (-0.02-0.06) |
| - follow up (~28 weeks) | -0.01 (-0.05-0.03) | -0.02 (-0.07-0.03) | -0.01 (-0.05-0.03) | 0.00 (-0.04-0.04) | -0.01 (-0.05-0.03) |
| MVV in litres per minute |  |  |  |  |  |
| - post-treatment (6 weeks) | 10.57 (3.26-17.88) | 14.3 (6.1-22.5) | 10.09 (2.11-18.07) | 10.32 (2.91-17.73) | 10.57 (3.3-17.85) |
| - follow up (~28 weeks) | 5.20 (-2.33-12.73) | 7.29 (-1.00-15.59) | 3.04 (-5.38-11.46) | 6.01 (-1.77-13.78) | 5.25 (-2.27-12.76) |
| PEF in litres per second |  |  |  |  |  |
| - post-treatment (6 weeks) | 0.38 (-0.24-1.00) | 0.52 (-0.17-1.22) | 0.35 (-0.29-0.99) | 0.35 (-0.29-0.99) | 0.38 (-0.24-1.00) |
| - follow up (~28 weeks) | -0.02 (-0.66-0.62) | -0.16 (-0.86-0.55) | -0.18 (-0.83-0.48) | 0.03 (-0.62-0.67) | -0.03 (-0.67-0.61) |
| *Quality of life* |  |  |  |  |  |
| SF-12 PCS |  |  |  |  |  |
| - post-treatment (6 weeks) | 3.79 (1.24-6.35) | 3.70 (0.76-6.63) | 3.68 (1.13-6.24) | 3.27 (0.69-5.86) | 3.75 (1.22-6.27) |
| - follow up (~28 weeks) | 2.69 (0.06-5.32) | 2.37 (-0.57-5.30) | 2.31 (-0.43-5.05) | 2.44 (-0.28-5.16) | 2.72 (0.12-5.33) |
| SF-12 MCS |  |  |  |  |  |
| - post-treatment (6 weeks) | 2.18 (-0.54-4.90) | 1.92 (-1.14-4.97) | 2.17 (-0.57-4.91) | 1.65 (-1.07-4.38) | 2.18 (-0.53-4.89) |
| - follow up (~28 weeks) | 1.99 (-0.81-4.79) | 1.48 (-1.58-4.54) | 2.30 (-0.48-5.09) | 1.82 (-0.96-4.61) | 1.93 (-0.87-4.73) |
| *mMRC dyspnoea, favourable outcome* |  |  |  |  |  |
| - interim 2 weeks | 1.27 (0.88-1.82) | 1.33 (0.90-1.98) | 1.26 (0.87-1.81) | 1.26 (0.88-1.82) | 1.28 (0.89-1.85) |
| - interim 4 weeks | 1.08 (0.82-1.42) | 1.04 (0.76-1.42) | 1.06 (0.80-1.4) | 1.06 (0.80-1.40) | 1.09 (0.82-1.43) |
| - post-treatment (6 weeks) | 1.46 (1.17-1.82) | 1.43 (1.14-1.80) | 1.42 (1.13-1.79) | 1.40 (1.11-1.77) | 1.47 (1.17-1.84) |
| - follow up (~28 weeks) | 1.22 (0.92-1.61) | 1.15 (0.84-1.57) | 1.21 (0.92-1.58) | 1.23 (0.92-1.63) | 1.23 (0.93-1.62) |

Apart from estimates for mMRC perceived dyspnoea (favourable outcome), estimates are between-group differences in mean change from baseline derived from linear mixed effects models adjusted for study centre (fixed effect) and with baseline means constrained to be equal across comparison groups. Estimates for mMRC perceived dyspnoea are rate ratios derived from general linear mixed model of the Poisson family with log link adjusted for centre and ln(number valid observations up to data point) as offset. CIs are estimated with cluster robust standard errors (cluster variable: participant ID).
\* Based on per protocol sample, i.e. participants who completed all assessments and completed the intervention program at or above minimum intensity and duration required for compliance as defined in the protocol (intervention group). <sup>§</sup> Based on multiple imputation using chained equations assuming data were missing at random. The imputation model included all outcomes and the following auxiliary variables with complete baseline information: gender, age, smoking history (no vs. yes), presence of any comorbidity (no vs. yes), Covid-19 severity (non-severe vs. severe), body mass index, time from admission to hospital to baseline assessment in days. \* Used reference-based controlled multi-variate normal imputation for each outcome assuming increments in both comparison groups followed the observed pattern of the control group where data points were not observed. Auxiliary variables employed were as in the MAR model. After imputation adaptive rounding was used for imputed values of mMRC dyspnoea (favourable outcome). <sup>§</sup> Same as main analysis but providing additional adjustment for time from onset of symptoms to measurement point in days and time from onset of symptoms to measurement point squared. Likelihood ratio test confirmed superior fit of model including squared term in addition.
CI = confidence interval, ITT = intention to treat, MAR = missing at random, CIR = copy increments from reference, 6MWD = six minute walking distance, FEV1 = forced expiratory volume in one second, FVC = forced vital capacity, MVV = maximum voluntary ventilation, PEF = peak expiratory flow, SF-12 = Medical Outcomes Short Form-12, PCS = physical component score, MCS = mental component score, mMRC = modified Medical Research Council.

### Adverse events

No serious adverse events occurred during the study period. Eight patients (5 from the TERECO group and 3 from the control group) were hospitalized, all for non-life-threatening reasons unrelated to Covid-19 or the intervention and all in the follow-up period. In total 174 adverse events occurred involving 73 (61.3%) patients (38 or 63.3 % in the control and 35 or 59.3% in the TERECO group). Of the events 78 were recorded in the control and 96 in the TERECO group. 64 events occurred during the intervention period (19 in controls and 45 in the TERECO group). All but one of those events (stomach ulcers of one patient in the control group were rated as moderately severe) were classified as very mild to moderate. These events mostly comprised uncomfortable symptoms including chest tightness, feelings of weakness or reduced physical strength, and cough. A detailed account of adverse events is provided in Appendix 3.

## DISCUSSION

In this trial, the TERECO program was superior to the control group regarding improvement of functional exercise capacity (6MWD), LMS (squat time), and the physical component of HRQOL (SF-12 PCS). All these effects could be sustained over a 7-months period. Pronounced differences in exercise capacity and lower-limb muscle strength remained between intervention and control group. For physical HRQOL, the difference between TERECO and the control group decreased at the final follow-up due to improvements in the control group. We also found a short-term effect of TERECO on MVV and mMRC perceived dyspnoea. Both effects, however, decreased at follow-up with differences from the control group no longer being statistically significant. No effects of TERECO on the four other pulmonary function parameters and statistically uncertain effects on the mental component of HRQOL were found in this study. Adherence to the intervention program was satisfactory and no serious adverse events occurred. All seven moderately-severe events that needed in-hospital treatment were unrelated to the intervention.

This study evaluated a relatively inexpensive, patient-centred, adaptable telerehabilitation intervention with a wide range of parameters of relevance to function and HRQOL of Covid-19 survivors including a comprehensive report on adverse events. With a few exceptions, this trial was executed according to the original protocol and the attrition rate was low (about 12%). Extensive sensitivity analysis was carried out that demonstrated the stability of most results under different scenarios including per protocol analysis as well as an extended MAR and a non-MAR scenario.

Limitations of this research include participant characteristics: only Covid-19 survivors with moderate dyspnoea symptoms who had previously been hospitalized for treatment were included. The results are thus not generalizable to persons with no, mild or severe dyspnoea, nor to people who contracted SARS CoV-2 but were not hospitalised for it. Another important weakness is the unexpected change of the location and resulting delay of the final follow-up assessment because regulations pertinent to the protection of health professionals from infection no longer permitted assessment visits. While conditions remained the same between intervention and control group, it is unclear how this may have affected patient-reported assessments and pulmonary function testing. Low-certainty evidence (one randomised crossover trial) suggests that 6MWT performed outdoors yields comparable results to centre-based testing.^40^ Emerging evidence suggest that most profound impairment of lung function in Covid-19 occurs in diffusion capacity^2 41^ usually measured as the diffusion capacity of the lung for carbon monoxide (CO) or the transfer capacity for CO. This was unclear at the time of study design and the required measurement procedures are difficult to perform in the home setting. Comparability of the occurrence of adverse events across groups during the intervention period was also limited by additional prompts in the intervention group issued in weekly consultations with therapists and through RehabApp. Recollection bias in the control group is possible, for instance, rather mild events that happened several days ago might not have been remembered during two-weekly phone interviews. Finally, this trial was not powered for subgroup analysis and effect sizes in specific subpopulations hence remain unclear.

At the time of writing, there are no other randomised-controlled studies on rehabilitation effectiveness for Covid-19. Demonstrating clinically meaningful and sustainable effects of the TERECO program on 6MWD and LMS, this study also adds to previous low-certainty evidence on the effectiveness of telerehabilitation in respiratory disease.^15^ The effect size for 6MWD in the present study at six weeks is comparable to results from a randomized-controlled trial from Hong Kong by Lau and associates^11^ who evaluated a 6-week outpatient exercise program for SARS survivors with baseline and post-treatment assessment. In contrast to our findings, the latter study did not detect any effects of the program on quality of life (SF-36) or lower-limb strength (measured as gluteus maximum and anterior deltoid strength with dynamometer). While a recent systematic review and meta-analysis reported superior effects of breathing exercise on lung function parameters (FEV1 and FEV1/FVC) as compared to control for COPD^42^, no such effects were found in the present study. In contrast to COPD, Covid-19 does not necessarily involve permanent impairment of structures of the lung and patients in our study from both groups seem to have naturally improved regarding lung function parameters. Another possible explanation is that the BCTE exercises included in the TERECO program were not sufficiently targeted. Moreover, compliance with this exercise type is difficult to measure remotely. Similar to the HRQOL physical component, mental aspects of quality of life also improved in both groups but no statistically significant between-group differences in increments were detected at any point, although the SF-12 MCS score of the TERECO group remained at about 2 points above control at follow-up. This result is difficult to interpret due to the unavailability of an MID for SF-12 in the target population. It is possible that our study was simply underpowered to detect a clinically relevant difference. In contrast to HRQOL, the proportion of patients free of subjective dyspnoea clearly decreased in the TERECO group between a peak at post-treatment and follow-up, returning to about the value at four weeks and suggesting that effects on perceived dyspnoea could not be sustained.

The TERECO program is targeted at improving physical fitness including physical aspects of subjective HRQOL and should be applied in populations with respective deficits. Effects of the program on pulmonary function are unlikely while those on mental well-being remain unclear. Components better targeting these outcomes could be added in future evaluations of similar programs. For example, low- to moderate-quality evidence for the effectiveness of Tai-Chi in improving lung function in persons with COPD exists.^43^ The TERECO program appears to be safe but more mild adverse events occurred in the TERECO group in the first 6 weeks. The occurrence of uncomfortable symptoms during and after exercise can possibly be reduced by adding more resting periods and warm up elements. Potential benefits of the latter should, however, be weighed against an increased amount of time needed for exercising. The TERECO program is inexpensive and suitable for large scale implementation dependent on smartphone coverage and the availability of therapists for remote supervision and consultations. The need for home visits or hospital attendance for program evaluation could be further reduced by the application of remote testing methods^40^.

## CONCLUSIONS

The TERECO program was superior over no rehabilitation with regard to functional exercise capacity, LMS, and the physical component of HRQOL. Only short-term effects were found for self-reported dyspnoea and MVV. Effects of the intervention on pulmonary function are otherwise unlikely and effects on mental aspects of quality of life are small at best.

## Supporting information

SAP

Marginal estimates from models

Distribution of adverse events

CONSORT checklist

## Data Availability

Anonymised patient-level data on which the analysis, results, and conclusions reported in this paper are based will be shared on a public digital repository upon publication of the paper.

## AUTHOR CONTRIBUTIONS

JL, WX, CZha, SL, and JDR designed the study with support from ZY, JW, YC, and CZhe; JL led the overall investigation and WX, CZha, SL, and YZ led the investigations at the respective centres; JL acquired the primary funding; WX, CZha, SL, JW, ZY, YC, CZhe, XF, and WC acquired and curated the data with JDR responsible for data checks and preparing the data for statistical analysis; JDR planned and performed the statistical analysis with support from SL and JW; JDR created all figures and tables for this article with support from JW; JDR wrote the original draft with support from SL and JW; all other authors revised the draft for critical content; JL and JDR supervised the study. All authors have read and agreed with the submitted version of the manuscript. JL and JDR act as guarantors for this work and accept full responsibility for the work and the conduct of the study, they had access to the data, and controlled the decision to publish.

## ACKNOWLEDGEMENTS

We sincerely appreciate the work of all our colleagues from The First Affiliated Hospital of Nanjing Medical University, Hubei Province Hospital of Integrated Chinese and Western Medicine, and Hubei Huangshi Hospital of Traditional Chinese Medicine who were involved in the conduct of this trial. Special thanks go to Dr. Yongqiang Li, Puyang Zhang, Wei Zhang, and Wenzhong Zou. We cordially thank all study participants and our other patients with Covid-19 for their feedback and ideas on how to design and improve the TERECO program. We thank Jiayi Zhang for cross-checking the English translation of the Chinese protocol. We like to thank our friend and linguistics expert Dr. Michael Herrera for his help with describing the different exercises. We are grateful to our friend Dr. Andrew ‘Randy’ Pennycott for review of style and language. JDR would like to thank Lili and Binhua Reinhardt for their patience and continuous support and Dietrich Reinhardt for excellent discussions about the lung function parameters and other aspects of data interpretation.

## DECLARATIONS AND STATEMENTS

### Ethical approval

This study was registered at the Chinese Clinical Trial Registry on 11 Apr 2020 (ChiCTR2000031834). Ethical approval was first received from the Institutional Review Board (IRB) of the First Affiliated Hospital of Nanjing Medical University/Jiangsu Province Hospital (2020-SR-171, 9 April 2020) and then subsequently from the IRBs of Hubei Province Hospital of Integrated Chinese and Western Medicine (2020016, 14 April 2020), and Huangshi Hospital of Chinese Medicine (HSZYPJ-2020-026-01, 20 April 2020).

### Declaration of interest

All authors have completed the ICMJE uniform disclosure form at www.icmje.org/coi_disclosure.pdf and declare: no support from any organisation for the submitted work. JL reports a research grant from Jiangsu CF PharmTech, Inc. JDR reports research grants from Xi’an Bureau of Technology, China; from the Fundamental Research Funds for the Central Universities, China; and from Sichuan Bureau of Science and Technology, China during the conduct of the study. No other relationships or activities exist that could appear to have influenced the submitted work.

### Funding

This research was funded by Jiangsu CF PharmTech, Inc (to JL); Department of Science and Technology of Sichuan Province (21ZDYF1918, to JDR), the Fundamental Research Funds of Central Universities in China (20827041D4161, to JDR), Xi’an Science and Technology Bureau (XA2020-HWYZ-0043, to JDR).

### Details of the role of the study sponsors

CF PharmTech, Inc provided technical and financial support for the conduct of the study. Department of Science and Technology of Sichuan Province, the Fundamental Research Funds of Central Universities in China, and Xi’an Science and Technology Bureau provided financial support for the conduct of the study.

### Statement of independence of researchers from funders

The funders did not have any influence on the design, conduct, analysis, data interpretation, and conclusions of his study.

### Transparency declaration

JL and JDR affirm that the manuscript is an honest, accurate, and transparent account of the study being reported; that no important aspects of the study have been omitted; and that any discrepancies from the study as planned (and, if relevant, registered) have been explained.

### Dissemination declaration

We have reported whether we plan to disseminate the results to study participants and or patient organisations or stated that dissemination to these groups is not possible/applicable.

---

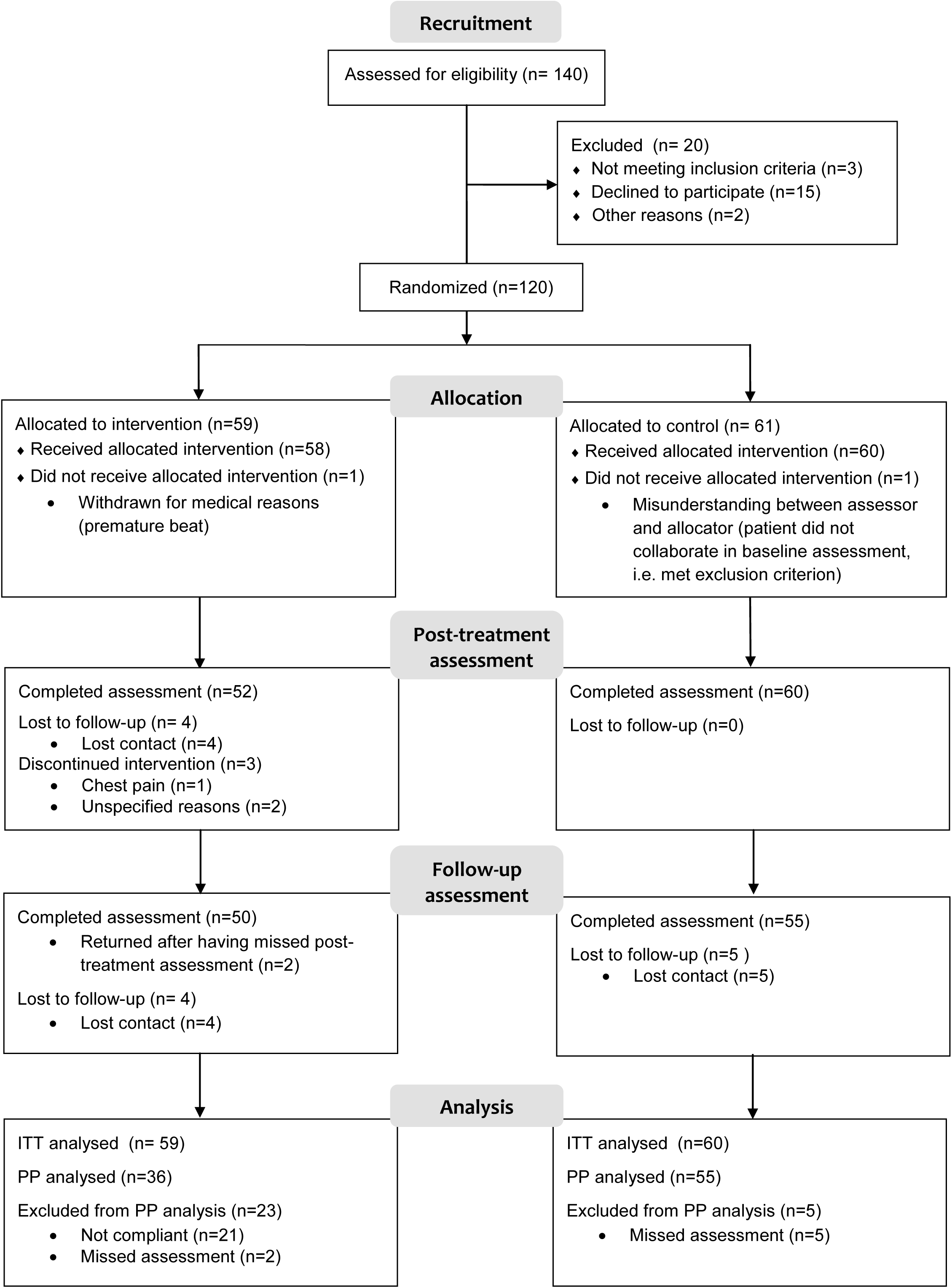

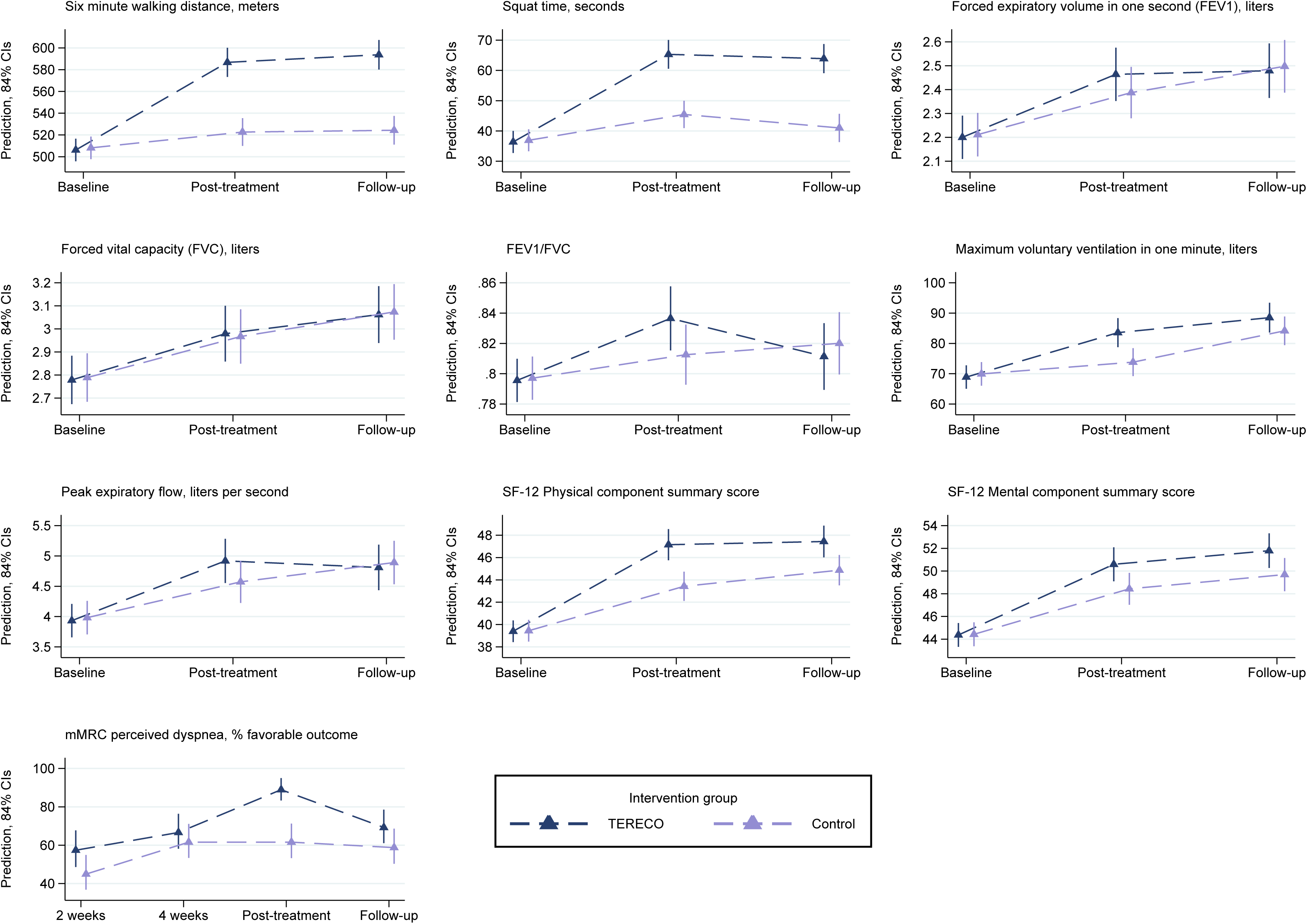

## Notes

### Competing Interest Statement

Jian'an Li has received a research grant from Jiangsu CF PharmTech, Inc. Jan D. Reinhardt reports research grants from Xi'an Bureau of Technology, China, from the Fundamental Research Funds for the Central Universities, China, and from Sichuan Bureau of Science and Technology, China during the conduct of the study. No other relationships or activities exist that could appear to have influenced the submitted work.

### Clinical Trial

ChiCTR2000031834

### Clinical Protocols

http://idmr.scu.edu.cn/info.htm?id=1841614474692833

