## Supplementary material for "Effectiveness of a telerehabilitation program for COVID-19 survivors (TERECO) on exercise capacity, pulmonary function, lower limb muscle strength, and quality of life: a randomized controlled trial": SAP

### Statistical Analysis Plan

### for

### Contents

I. Administrative information ........ - 3 -
II. Introduction ........ - 4 -
III. Study methods ........ - 5 -
IV. Statistical principles ........ - 6 -
V. Trial Population ........ - 7 -
VI. Analysis ........ - 9 -
VII. References ........ - 13 -

### I. ADMINISTRATIVE INFORMATION

**Study title:** Effectiveness of a home-based and remotely supervised telerehabilitation program for COVID-19 survivors (TERECO): randomized controlled trial

**Principal Investigator:** Jianan Li, MD

**Senior statistician:** Jan D. Reinhardt, PhD

**Trial registry/Number/Date:** Chinese Clinical Trial Registry/ChiCTR2000031834/11 Apr 2020

**Version of protocol/Date**: V1.9/20 Apr 2020

**Version of SAP/Date**: V1/20 Apr 2020 (version used for protocol V1.9)

**Contributors**: Jan D. Reinhardt (lead author, senior statistician), Jiayue Wang (contributing author, translation to Chinese for protocol), Shouguo Liu (contributing author), Jianan Li (principle investigator)

**Affiliations**: (1) Sichuan University, Chengdu, China – Jan D. Reinhardt; (2) Swiss Paraplegic Research, Nottwil Switzerland – Jan D. Reinhardt (3) Department of Health Sciences and Medicine, University of Lucerne, Switzerland – Jan D. Reinhardt. (4) Center of Rehabilitation Medicine, The First Affiliated Hospital of Nanjing Medical University, Nanjing, China – Jiayue Wang, Shouguo Liu, and Jianan Li; (2) School of Rehabilitation Medicine, Nanjing Medical University, Nanjing, Jiangsu, China – Jiayue Wang, Shouguo Liu, and Jianan Li

**Contact**: Prof. Dr. Jan D. Reinhardt, Institute for Disaster Management and Reconstruction of Sichuan University and Hong Kong Polytechnic University, 122 Huang He Zhong Lu (first section), 610207 Chengdu, Sichuan, China,; phone: +86-159-28554027

### II. INTRODUCTION

#### 2.1 Trial background

After discharge from acute treatment, a considerable number of patients with coronavirus disease-2019 (COVID-19) will likely experience longer-term health problems and disability including reduced exercise capacity, impairment of lung function, limitations in activities of daily living, and reduced quality of life. Addressing those rehabilitation needs with conventional in- or outpatient pulmonary rehabilitation programs(1-3) is limited however due to the ongoing Covid-19 pandemic and associated risk of disease transmission as well as lockdown measures currently employed in China. Remotely supervised telerehabilitation programs provide a viable alternative(4). The telerehabilitation program for COVID-19 survivors (TERECO) is an exercise-based pulmonary rehabilitation program developed according to the standards of the Chinese Association of Rehabilitation Medicine(5) that combines a smart-phone application, remote monitoring techniques, and tele-consultations with clinicians in order to provide a low-cost, safe, effective, and easy-to-operate means for patients to recover at home.

#### 2.2 Trial objectives

The primary objective of the planned randomized controlled trial evaluate the immediate (post-intervention) and sustained (longer-term) effectiveness of the TERECO program on exercise capacity, pulmonary function, dyspnea, lower limb muscle strength, and quality of life in patients with coronavirus disease 2019 who have been discharged from hospital.

Additional objectives are to monitor the occurrence of adverse events in both groups and to evaluate the feasibility of TERECO in terms of design (e.g. appropriateness of exercise specifications), safety (unintended side effects) and compliance.

### III. STUDY METHODS

#### 3.1 Design

Pragmatic, multi-center, randomized-controlled, 1:1 parallel group trial. The control group receives educational instructions for daily living at baseline. The intervention group participates in a 6-weeks, home-based, remotely monitored exercise program for cardiopulmonary rehabilitation, delivered via smartphone (R+ health app software) with 3-4 sessions per week and monitored remotely in real-time via a heart rate device worn on the body of the patient. Exercises types include breathing control and thoracic expansion, aerobic exercise, and muscle strength exercises. Exercise difficulty, intensity, and duration is scheduled to increase over time.

#### 3.2 Randomization and allocation concealment

Permutated block-randomization stratified by study center (block size permuted in the range of 10-14) with allocation concealed by central randomization.

#### 3.3 Sample size calculation

Sample size is calculated for the primary outcome 6 minute walking distance (6MWD) measured with 6 minutes walking test (6MWT). A systematic review on measurement properties of field walking tests conducted by the European Respiratory Society and American Thoracic Society(6) found a median minimal important difference (MID) of 30 meters for the 6MWT (range 22-57 meters) across studies using anchor- or distribution-based approaches in different lung disease including COPD, pulmonary arterial hypertension, and interstitial lung disease. Since an MID for 6MWT in COVID-19 has not been established at the time of the development of the TERECO protocol and learning effects may also occur (particularly in the exercise group), a larger and thus more conservative MID of 50 meters is supposed here. A standard deviation of change in 6MWD over six weeks of 99 meters in controls and 71 in the intervention group as reported in a previous trial on the effectiveness of an exercise program in SARS survivors(3) is further assumed. The assumption of a greater standard deviation in controls makes sense in so far as a greater variety of outcomes can be expected in this group due to greater variations in disease course and environmental influences as these are not influenced by a targeted intervention. With a power of 80% and an alpha error of 5% this yields a sample size of 96 participants respectively 48 per group to detect a statistically significant signal for a difference in change in 6MWD between groups with an independent samples t-test. This corresponds to power analysis for ANCOVA or constrained longitudinal data analysis for high correlations ($r>=0.8$) between repeated measurements(7, 8). An assumption of such high correlation is conservative and justified given that we have no previous knowledge about the effectiveness of similar interventions in the target population. Assuming an attrition rate of 20% the recruitment target is 120 participants or about 60 per group.

##### 3.4 Framework

Superiority of intervention over control.

##### 3.5 Statistical interim analysis and stopping guidelines

No formal interim analysis will be performed. If serious adverse events (death; or re-hospitalization related to the intervention or the studied disease) or uncontrolled disease progression occur in 10 percent or more of the study participants, the study will be terminated.

##### 3.6 Timing of analysis

Data analysis will begin after follow up assessments have been completed or patients are considered lost to follow up for all study participants. All outcomes will be collectively analyzed.

##### 3.7 Timing of outcome assessments

Outcomes will be assessed at visits to the patients home. Main assessments are planned at three time points. (1) A baseline assessment will be conducted after recruitment of the patient and signing of written informed consent but before group allocation is revealed. (2) A post-treatment assessment will take place after completion of the exercise program at around 6 weeks after baseline. (3) A follow up assessment will take place at around 24 weeks after baseline. Appointments for assessment visits at the patients' homes will be made.

#### IV. STATISTICAL PRINCIPLES

##### 4.1 Confidence intervals and P values

Level of statistical significance is set at alpha error = 0.05. No adjustments for multiplicity are made. Treatment effects will be reported with 95 % confidence intervals.

##### 4.2 Adherence and protocol deviations

***Adherence to intervention***

Total exercise time for a given day will be determined as the time from opening to closing the smartphone rehabilitation application as recorded by RehabApp. The time during aerobic exercise within which the monitored heart rate reaches the target heart rate or above will be counted as effective exercise time. Having reached at least two thirds (66.66%) of the scheduled total AND effective target time in any given week for at least five of the six weeks according to RehabApp records will be considered compliant with the protocol. Number and percentage of participants in the intervention group who adhered to the intervention program as defined above will be reported.

***Protocol deviations***

Any deviations from the protocol (v1.9, 20 April 2020) with regard to sample size, trial eligibility, randomization and allocation, intervention delivery, outcome definitions and assessments, and the statistical analysis plan will be reported.

##### 4.3 Analysis population

Main analysis will be based on intention to treat including all available observations from participants as randomized. Analysis of the per protocol sample will be part of sensitivity analyses.

#### V. TRIAL POPULATION

##### 5.1 Eligibility

Inclusion criteria are

(1) COVID-19 (confirmed SARS CoV-2 infection);

(2) discharged from hospital-based acute treatment for Covid-19;

(3) moderate dyspnea symptom (mMRC 2~3);

(4) aged 18 to 75 years;

(5) possession of and ability to use smart phone (study subjects or family members);

(6) provided informed consent and signed the informed consent form.

Exclusion criteria are

(1) resting heart rate>100bpm;

(2) taking drugs which affect cardiopulmonary function or heart rate (such as: trimetazidine, salbutamol, beta-blocker);

(3) combined uncontrolled chronic disease, such as uncontrolled hypertension (resting BP≥160/100mmHg), uncontrolled diabetes (random blood glucose>16.7 mmol/l, HbA1C>7.0%);

(4) combined severe organic disease, such as unstable hemodynamic heart disease, heart failure with limited movement (previous diagnosis, or having clinical symptoms of heart failure, after consultation by cardiologist), unstable angina, MI within past 12 months or other cardiac diseases;

(5) cerebrovascular disease that occurred within the past 6 months;

(6) active stage of digestive ulcer, thyroid dysfunction, or active tuberculosis;

(7) chronic kidney disease >=stage 3 (eGFR<60ml/min);

(8) having received intra-articular drug injection or surgical treatment in lower extremities within past 6 months;

(9) poor compliance, unable to cooperate with assessment or training; (10) unable to walk independently with auxiliary;

(11) combined mental disease that prevents patients from living independently or receiving treatment;

(12) alcohol abuse or illegal drug use history;

(13) pregnant, nursing or preparing for pregnancy (including male subjects);

(14) having participated in other clinical trials within the past 3 months or currently enrolled in other clinical trial;

(15) enrolled in other rehabilitation program.

**5.2 Recruitment, withdrawal and follow up**

Hospital records will be pre-screened for eligibility. Patients will then be contacted by telephone and the study will be explained to them. If patients are interested in participation, a home visit for further evaluation of eligibility, signing of informed consent form, and baseline assessment will be scheduled.

If conditions occur for a study participant that make it unsuitable to continue the study, including: exacerbation of disease, serious adverse events, safety concerns regarding performance of exercise, new development of a health condition or symptoms listed under exclusion criteria, withdrawal of consent, poor compliance with assessments, etc., this patient will be withdrawn from the study.

Information to be included in the CONSORT flow diagram will be number screened for eligibility and number excluded with reasons, number randomized and number allocated to intervention for each trial arm, number who received the intervention as allocated by trial arm, number who completed the assessment and number who discontinued the intervention, were withdrawn for medical or other reasons, or were otherwise lost to follow up with reasons by trial arm and for each assessment point.

##### 5.3 Baseline patient characteristics

The following data will be extracted from patient records and confirmed upon the baseline visit where applicable: age, gender, occupation at disease onset, height, weight, presence of comorbidities, disease severity, treatment modalities during hospitalization for Covid-19, admission and discharge dates.

Demographic and relevant clinical baseline data will be expressed by descriptive statistics (number and proportion or mean ± standard deviation, in total and by intervention group).

#### VI. ANALYSIS

##### 6.1 Outcome definitions

***Primary outcome:*** The primary outcome of this trial is change in distance walked in meters within six minutes (6MWD) from baseline to 6 weeks (end of intervention program) assessed with the 6-minutes walking test (6MWT).(6, 9)

***Secondary outcomes***

(1) Change in 6MWD in meters from baseline to 24 weeks.

(2) Pulmonary function assessed with a portable pulmonary function device according to the guidelines of the American Thoracic Society.(10, 11) Parameters to be measured include forced

vital capacity (FVC), forced expiratory volume in the first second (FEV1), the ratio of FEV1 and FVC (FEV1/FVC), Peak Exploratory Flow (PEF), and Maximum Voluntary Ventilation (MVV). Percent of predicted value for FEV1, FVC, and FEV1/FVC will be calculated based on Global Lung Initiative (GLI-2012) equations for South-East Asia(12). Percent of predicted PEF and MVV will be calculated based on equations for mainland China provided by Mu and Liu(13).

(3) Perceived dyspnea assessed with the modified Medical Research Council scale (mMRC)(14). This is a patient-reported ordinal scale which distinguishes five grades of dyspnea with 0 indicating no dyspnea and 5 indicating extreme problems with breathlessness making it impossible to leave the house or occurring while dressing or undressing. Given that patients will be included in this trial only if they have baseline mMRC scores of 2-3, an mMRC score of zero is defined as a favorable outcome.

(4) Lower limb muscle strength and endurance measured with the static squat test(15). Participants are asked to perform a squat with their back against the wall starting from a standing position, feet flat on the ground, and approximating a 90° angle at the hip and knees. The time in seconds participants can remain in this squatting positions will be recorded.

(5) Quality of life evaluated with Short Form Health Survey-12 (SF-12) Version 2 (16, 17) reported as two composite scores: Physical Health and Mental Health Composite score, with higher score indicating better health. Scores will be standardized according to US norms(17, 18) for reasons of international comparability and due to lack of general population reference data for mainland China.

##### 6.2 Analysis methods

Main analysis of treatment effects will be performed on an intention to treat basis. Crude change in outcomes for both groups from baseline to post-treatment and follow up will be provided as mean change scores with standard deviations or proportions.

***Main analysis of primary outcome***

The primary outcome will be analyzed with constrained longitudinal data analysis (cLDA), i.e. a linear mixed effects model with a random intercept for subject will be fitted to regress 6MWD at baseline and 6 weeks on study center, time point of assessment and an interaction of time point and intervention group.(7, 19) A mere group effect as in regular LDA such as repeated measures ANOVA is not included thus constraining baseline means for groups to be equal. This type of model adjusts for baseline mean differences in the outcome between intervention groups similar to analysis of covariance (ANCOVA). In fact, it is equivalent to ANCOVA in the absence of missing outcome data. In the presence of missing data cLDA in contrast to ANCOVA considers all available data in the estimation of the common baseline mean and avoids listwise deletion of cases lost to follow up.

***Main analysis of secondary outcomes***

For analysis of the overall trajectory of 6MWD until final follow at 24 weeks, data from all time points will be considered and terms for the 24 week time point and its interaction with intervention group will be added to the above model. Secondary continuous outcomes (pulmonary function parameters, lower limb muscle strength as time in seconds remaining in squatting position, and SF-12 composite scores) will be analyzed analogously, estimating treatment effects at 6 weeks and 24 weeks simultaneously. Occurrence of a favorable outcome with regard to dyspnea (mMRC) will be analyzed with longitudinal logistic regression(20). Estimates of treatment effects and changes in mean scores (continuous outcomes) or percentages (discrete outcomes) from baseline to 6 weeks and 24 weeks follow up by intervention group will be provided with 95% confidence intervals. Residuals of all linear models will be checked for normal distribution using qq-plots; if this assumption is violated robust standard errors (Huber-White sandwich estimator) will be calculated.

***Adjustments***

All main analysis of treatment effects will be adjusted for study center (as fixed effect).

***Corrections for multiplicity***

No corrections for multiple testing will be applied.

***Missing data***

As mixed effects models are used for analysis of treatment effects and listwise deletion of cases with missing data is thus avoided, main analysis will be performed on available (unimputed) data, assuming that data are missing at random (MAR) and missing values depend on observed variables in the model (center, time point, and the interaction of group and time point)(21, 22). This assumption will be tested in sensitivity analysis (see below).

***Sensitivity analysis and per protocol analysis***

Two types of sensitivity analysis will be performed. First, above analyses will be repeated on 50 multiply imputed datasets(23) whereas imputation is based on available outcome data as well as available baseline information including variables not included in the analysis models. This compares the simple MAR assumption outlined above with an extended MAR assumption, i.e. that missing values are also dependent on observed (auxiliary) variables not included in the models used for primary analysis. Apart from center, these auxiliary variables comprise gender, age, disease severity, time from first hospital admission for Covid-19 to baseline assessment, presence of comorbidities, smoking history, and body mass index. Imputation is preferred over adjustment within the mixed models (adding covariates) as conditioning on covariates is not intended given this is a pragmatic trial and to avoid overfitting of the models. Chained equations will be used for simultaneous imputation of all missing outcome data. Second, reference-based multiple imputation will be used for simulating a non-missing at random (NMAR) scenario where patients with missing assessments in the intervention group are supposed to experience change in the outcome that is similar to that observed in the control group after the last available data point before dropout. This is called copy increments in reference (CIR) approach. Imputation is based on a joint multivariate normal distribution of the outcome for each treatment arm. In the case of CIR those in the intervention group are supposed to switch to the distribution of the control group after dropout(24). CIR imputation is done separately for each outcome and considering center and auxiliary covariates specified for extended MAR imputation. Per protocol analysis will also be performed estimating the previously specified models on data including only those participants from the intervention group who had adhered to the TERECO protocol as defined above.

***Sub-group analysis***

Due to lack of power for this type of analysis, no pre-specified sub-group analysis will be performed.

***Analysis of harms***

Patients in the intervention group can actively report adverse events to the responsible doctor at any time through the smart phone app or contact the responsible doctor, clinical department, or researcher by phone or WeChat. Patients in the control group can actively report adverse events by contacting the responsible doctor, the clinical department, or the researcher by phone or WeChat. In addition, all patients will be asked at 2 and 4 weeks telephone consultations, at the 6 weeks post-treatment assessment and at 24 weeks follow-up assessment if any adverse events occurred during the previous period. Severity and relationship with intervention of these adverse events will be evaluated by two independent doctors otherwise uninvolved in the trial. Conflicts will be resolved by consensus or if not possible by consultation of a third reviewer.

Proportions of patients reporting any adverse events and serious adverse events during the whole study period, during the intervention period, and during the follow up period stratified by intervention group as randomized will be reported. Number of adverse events per patient reporting any such events will also be given as well as types/severity of events and supposed relationship with interventions.

***Software***

All analyses will be performed with Stata 14 (Stata Corporation, Texas, USA).
