## Supplementary material for "Effectiveness of a telerehabilitation program for COVID-19 survivors (TERECO) on exercise capacity, pulmonary function, lower limb muscle strength, and quality of life: a randomized controlled trial": Marginal estimates from models

**Appendix 3: Marginal estimates for outcomes across time points and groups with 84% confidence intervals, equality constraints imposed for baseline means of continuous outcomes and adjusted for center**

| Physical performance measures | | | | |
|---|---|---|---|---|
| | Estimated margin | SE | 84% CI | |
| ***6MWD in meters*** | | | | |
| *Baseline* | | | | |
| TERECO | 506.20 | 7.37 | 495.84 | 516.56 |
| Control | 508.16 | 7.37 | 497.80 | 518.51 |
| *Post-treatment* | | | | |
| TERECO | 586.74 | 9.53 | 573.35 | 600.13 |
| Control | 522.68 | 9.08 | 509.93 | 535.44 |
| *Follow-up* | | | | |
| TERECO | 593.71 | 9.65 | 580.15 | 607.27 |
| Control | 524.29 | 9.38 | 511.11 | 537.46 |
| ***Squat time in seconds*** | | | | |
| *Baseline* | | | | |
| TERECO | 36.37 | 2.58 | 32.75 | 40.00 |
| Control | 36.93 | 2.58 | 33.30 | 40.56 |
| *Post-treatment* | | | | |
| TERECO | 65.31 | 3.37 | 60.57 | 70.05 |
| Control | 45.47 | 3.21 | 40.96 | 49.98 |
| *Follow-up* | | | | |
| TERECO | 63.90 | 3.42 | 59.10 | 68.70 |
| Control | 41.00 | 3.31 | 36.35 | 45.66 |

| Spirometry | | | | |
|---|---|---|---|---|
| | Estimated margin | SE | 84% CI | |
| ***FEV1 in liters*** | | | | |
| *Baseline* | | | | |
| TERECO | 2.20 | 0.06 | 2.11 | 2.29 |
| Control | 2.21 | 0.06 | 2.12 | 2.30 |
| *Post-treatment* | | | | |
| TERECO | 2.46 | 0.08 | 2.35 | 2.58 |

| | | | | |
|---|---|---|---|---|
| Control | 2.39 | 0.08 | 2.28 | 2.49 |
| *Follow-up* | | | | |
| TERECO | 2.48 | 0.08 | 2.37 | 2.59 |
| Control | 2.50 | 0.08 | 2.39 | 2.61 |
| ***FVC in liters*** | | | | |
| *Baseline* | | | | |
| TERECO | 2.78 | 0.07 | 2.67 | 2.88 |
| Control | 2.79 | 0.07 | 2.68 | 2.89 |
| *Post-treatment* | | | | |
| TERECO | 2.98 | 0.09 | 2.86 | 3.10 |
| Control | 2.97 | 0.08 | 2.85 | 3.08 |
| *Follow-up* | | | | |
| TERECO | 3.06 | 0.09 | 2.94 | 3.19 |
| Control | 3.07 | 0.09 | 2.95 | 3.19 |
| ***FEV1/FVC*** | | | | |
| *Baseline* | | | | |
| TERECO | 0.80 | 0.01 | 0.78 | 0.81 |
| Control | 0.80 | 0.01 | 0.78 | 0.81 |
| *Post-treatment* | | | | |
| TERECO | 0.84 | 0.02 | 0.82 | 0.86 |
| Control | 0.81 | 0.01 | 0.79 | 0.83 |
| *Follow-up* | | | | |
| TERECO | 0.81 | 0.02 | 0.79 | 0.83 |
| Control | 0.82 | 0.01 | 0.80 | 0.84 |
| ***MVV in liters per minute*** | | | | |
| *Baseline* | | | | |
| TERECO | 68.90 | 2.75 | 65.04 | 72.76 |
| Control | 69.95 | 2.76 | 66.07 | 73.83 |
| *Post-treatment* | | | | |
| TERECO | 83.57 | 3.40 | 78.78 | 88.35 |
| Control | 73.84 | 3.28 | 69.24 | 78.44 |
| *Follow-up* | | | | |
| TERECO | 88.51 | 3.49 | 83.62 | 93.41 |
| Control | 84.17 | 3.36 | 79.45 | 88.89 |
| ***PEF in liters per second*** | | | | |
| *Baseline* | | | | |

| | | | | |
|---|---|---|---|---|
| TERECO | 3.93 | 0.20 | 3.66 | 4.21 |
| Control | 3.98 | 0.20 | 3.71 | 4.26 |
| *Post-treatment* | | | | |
| TERECO | 4.92 | 0.26 | 4.55 | 5.28 |
| Control | 4.57 | 0.25 | 4.23 | 4.92 |
| *Follow-up* | | | | |
| TERECO | 4.81 | 0.27 | 4.44 | 5.19 |
| Control | 4.89 | 0.25 | 4.53 | 5.25 |
| | **Quality of life** | | | |
| | Estimated margin | SE | 84% CI | |
| ***SF-12 PCS*** | | | | |
| *Baseline* | | | | |
| TERECO | 39.40 | 0.69 | 38.43 | 40.37 |
| Control | 39.45 | 0.69 | 38.48 | 40.42 |
| *Post-treatment* | | | | |
| TERECO | 47.15 | 0.99 | 45.75 | 48.55 |
| Control | 43.43 | 0.93 | 42.12 | 44.74 |
| *Follow-up* | | | | |
| TERECO | 47.44 | 1.01 | 46.02 | 48.86 |
| Control | 44.87 | 0.97 | 43.51 | 46.23 |
| ***SF-12 MCS*** | | | | |
| *Baseline* | | | | |
| TERECO | 44.37 | 0.75 | 43.32 | 45.42 |
| Control | 44.43 | 0.75 | 43.38 | 45.48 |
| *Post-treatment* | | | | |
| TERECO | 50.60 | 1.07 | 49.10 | 52.09 |
| Control | 48.43 | 1.00 | 47.03 | 49.84 |
| *Follow-up* | | | | |
| TERECO | 51.79 | 1.08 | 50.27 | 53.32 |
| Control | 49.69 | 1.04 | 48.23 | 51.15 |
| **mMRC perceived Dyspnea, favorable outcome in percent** | | | | |
| | Percent | SE | 84% logistic CI | |
| *2 weeks* | | | | |

| | | | | |
|---|---|---|---|---|
| TERECO | 57.38 | 0.12 | 48.63 | 67.70 |
| Control | 44.94 | 0.14 | 36.79 | 54.90 |
| *4 weeks* | | | | |
| TERECO | 66.63 | 0.10 | 58.13 | 76.38 |
| Control | 61.59 | 0.10 | 53.37 | 71.06 |
| *Post-treatment* | | | | |
| Intervention | 88.94 | 0.05 | 83.30 | 94.97 |
| Control | 61.59 | 0.10 | 53.26 | 71.21 |
| *Follow-up* | | | | |
| TERECO | 69.26 | 0.09 | 61.08 | 78.53 |
| Control | 58.76 | 0.11 | 50.33 | 68.60 |

All estimates apart from those mMRC perceived dyspnea are from constrained linear mixed effects models with random intercept for study participants, with equality constraint for baseline means and adjusted for center. Estimates for mMRC perceived dyspnea are from a generalized linear model from the Poisson family with a log link adjusted for center and ln(number of observations until data point) as offset, and using cluster-robust standard errors (cluster variable: participant ID). Baseline is zero percent favorable outcome for both groups and thus not estimated.

84% confidence intervals are used as comparison bars for visual inspection of margins in Figure 2. Overlap or non-overlap of 84% confidence intervals provides an approximation for comparing mean differences at alpha = 0.05.

SE = standard error, CI = confidence interval, TERECO = telerehabilitation intervention for Covid-19 survivors, 6MWD = six minute walking distance, FEV1 = forced expiratory volume in 1 second, FVC = forced vital capacity, MVV = maximum voluntary ventilation, PEF = peak expiratory flow, SF-12 = Medical Outcomes Short Form-12, PCS = physical component score, MCS = mental component score, mMRC = modified Medical Research Council
