## Supplementary material for "Effectiveness of a telerehabilitation program for COVID-19 survivors (TERECO) on exercise capacity, pulmonary function, lower limb muscle strength, and quality of life: a randomized controlled trial": Distribution of adverse events

**Appendix 5: Distribution of adverse events across study period and comparison groups**

| | Whole study period | | | During intervention period | | | During follow up period | | |
|---|---|---|---|---|---|---|---|---|---|
| | Total (n= 119) | Control (n=60) | TERECO (n=59) | Total (n= 119) | Control (n=60) | TERECO (n=59) | Total (n= 105) | Control (n=55) | TERECO (n=50) |
| **Per study participant** | | | | | | | | | |
| *Occurrence of any event, n(percent)* | | | | | | | | | |
| no | 46 (38.66) | 22 (36.67) | 24 (40.68) | 85 (71.43) | 51 (85.00) | 34 (57.63) | 45 (42.86) | 19 (34.55) | 26 (52.00) |
| yes | 73 (61.34) | 38 (63.33) | 35 (59.32) | 34 (28.57) | 9 (15.00) | 25 (42.37) | 60 (57.14) | 36 (65.45) | 24 (48.00) |
| *Number if occurred, n(percent)* | | | | | | | | | |
| 1 event | 29 (39.73) | 18 (47.37) | 11 (31.43) | 14 (41.18) | 2 (22.22) | 12 (48.00) | 31 (51.67) | 20 (55.56) | 11 (45.83) |
| 2 events | 20 (27.40) | 11 (28.95) | 9 (25.71) | 13 (38.24) | 5 (55.56) | 8 (32.00) | 18 (30) | 12 (33.33) | 6 (25.00) |
| 3 events | 9 (12.33) | 3 (7.89) | 6 (17.14) | 4 (11.76) | 1 (11.11) | 3 (12.00) | 5 (8.33) | 1 (2.78) | 4 (16.67) |
| more than 3 events | 15 (20.55) | 6 (15.79) | 9 (25.71) | 3 (8.82) | 1 (11.11) | 2 (8.00) | 6 (10) | 3 (8.33) | 3 (12.50) |
| *Hospitalized, n(percent)* | | | | | | | | | |
| no | 111 (94.12) | 57(95.00) | 54 (91.53) | 0 (0.00) | 0 (0.00) | 0 (0.00) | 97 (92.38) | 57(94.55) | 45 (90.00) |
| yes | 8 (6.72) | 3 (.005) | 5 (8.47) | 0 (0.00) | 0 (0.00) | 0 (0.00) | 8 (7.62) | 3 (5.45) | 5 (10.00) |
| **Per event** | | | | | | | | | |
| *Total number of events* | 174 | 78 | 96 | 64 | 19 | 45 | 110 | 59 | 51 |
| *Severity, n(percent)*[§] | | | | | | | | | |
| very mild, no follow up | 21 (12.07) | 8 (10.26) | 13 (13.54) | 12 (18.75) | 2 (10.53) | 10 (22.22) | 9 (8.18) | 6 (10.17) | 3 (5.88) |
| mild, further observation | 122 (70.11) | 61 (78.21) | 61 (63.54) | 34 (53.13) | 13 (68.42) | 21 (46.67) | 88 (80) | 48 (81.36) | 40 (78.43) |
| moderate, needs medical attention | 24 (13.79) | 4 (5.13) | 20 (20.83) | 17 (26.56) | 3 (15.79) | 14 (31.11) | 7 (6.36) | 1 (1.69) | 6 (11.76) |
| moderate to severe, needs treatment | 7 (4.02) | 5 (6.41) | 2 (2.08) | 1 (1.56) | 1 (5.26) | 0 (0) | 6 (5.45) | 4 (6.78) | 2 (3.92) |
| *Relationship with intervention,* | | | | | | | | | |

| | | | | | | | | | |
|---|---|---|---|---|---|---|---|---|---|
| *n(percent)*§ | | | | | | | | | |
| unlikely | 117 (67.24) | 62 (79.49) | 55 (57.29) | 10 (15.63) | 5 (26.32) | 5 (11.11) | 107 (97.27) | 57 (96.61) | 50 (98.04) |
| possible | 29 (16.67) | 10 (12.82) | 19 (19.79) | 27 (42.19) | 8 (42.11) | 19 (42.22) | 2 (1.82) | 2 (3.39) | 0 (0.00) |
| likely | 28 (16.09) | 6 (7.69) | 22 (22.92) | 27 (42.19) | 6 (31.58) | 21 (46.67) | 1 (0.91) | 0 (0.00) | 1 (1.96) |
| *Types of adverse events‡, n(percent)* | | | | | | | | | |
| chest tightness | 36 (20.69) | 13 (16.67) | 23 (23.96) | 10 (15.63) | 0 (0.00) | 10 (22.22) | 26 (23.64) | 13 (22.03) | 13 (25.49) |
| weakness | 18 (10.34) | 12 (15.38) | 6 (6.25) | 3 (4.69) | 2 (10.53) | 1 (2.22) | 15 (13.64) | 10 (16.95) | 5 (9.80) |
| cough | 14 (8.05) | 8 (10.26) | 6 (6.25) | 1 (1.56) | 0 (0.00) | 1 (2.22) | 13 (11.82) | 8 (13.56) | 5 (9.80) |
| reduced physical strength | 12 (6.9) | 6 (7.69) | 6 (6.25) | 0 (0.00) | 0 (0.00) | 0 (0.00) | 12 (10.91) | 6 (10.17) | 6 (11.76) |
| sputum discharge | 10 (5.75) | 5 (6.41) | 5 (5.21) | 0 (0.00) | 0 (0.00) | 0 (0.00) | 10 (9.09) | 5 (8.47) | 5 (9.80) |
| dizziness | 7 (4.02) | 1 (1.28) | 6 (6.25) | 7 (10.94) | 1 (5.26) | 6 (13.33) | 0 (.000) | 0 (0.00) | 0 (0.00) |
| chest pain | 6 (3.45) | 1 (1.28) | 5 (5.21) | 5 (7.81) | 0 (0.00) | 5 (11.11) | 1 (0.91) | 1 (1.69) | 0 (0.00) |
| back pain | 6 (3.45) | 3 (3.85) | 3 (3.13) | 5 (7.81) | 2 (10.53) | 3 (6.67) | 1 (0.91) | 1 (1.69) | 0 (0.00) |
| other | 65 (37.36) | 29 (37.18) | 36 (37.5) | 33 (51.56) | 14 (73.68) | 19 (42.22) | 32 (29.09) | 15 (25.42) | 17 (33.33) |

§ Rated by two doctors who had access to event history and medical history of each patient but were blinded to group allocation. Doctors were not otherwise involved in the study. Ratings were compared after a first round of independent rating (average ICC = 0.63 for severity ratings and 0.82 for relationship with intervention). Disagreement on particular events was resolved by discussion and consensus. If consensus could not be achieved a third doctor was consulted who had the casting vote. ‡ Listed are all adverse events with occurrence ≥ 5 events, all other events are subsumed under "other".
